# Antibiotics and hospital-associated *Clostridioides difficile* infection: systematic review and meta-analysis 2020 update

**DOI:** 10.1101/2021.02.21.21252172

**Authors:** Claudia Slimings, Thomas V. Riley

## Abstract

**Background:** *Clostridioides difficile* infection (CDI) is the most common cause of healthcare facility-associated (HCFA) infectious diarrhoea in high-income countries. Antibiotic use is the most important modifiable risk factor for CDI. The most recent systematic review covered studies published until 31^st^ December 2012.

**Objectives:** To update the evidence for epidemiological associations between specific antibiotic classes and HCFA-CDI for the period 1^st^ January 2013 to 31^st^ December 2020.

**Data sources:** PubMed, Scopus, Web of Science Core Collection, WorldCat, and Proquest Dissertations and Theses.

**Study eligibility criteria, participants and exposures:** Eligible studies were those conducted among adult hospital inpatients, measured exposure to individual antibiotics or antibiotic classes, included a comparison group, and measured the occurrence of HCFA-CDI as an outcome.

**Study appraisal and synthesis methods:** The Newcastle–Ottawa Scale for the Assessment of Quality was used to appraise study quality. To assess the association between each antibiotic class and HA-CDI, a pooled random effects meta-analysis was undertaken. Metaregression and sub-group analysis was used to investigate study characteristics identified *a priori* as potential sources of heterogeneity.

**Results:** Carbapenems, and 3^rd^ and 4^th^ generation cephalosporin antibiotics remain most strongly associated with HCFA-CDI, with cases more than twice as likely to have recent exposure to these antibiotics prior to developing CDI. Modest associations were observed for fluoroquinolones clindamycin, and beta-lactamase inhibitor combination penicillin antibiotics.

**Limitations:** Individual study effect sizes were variable and heterogeneity was observed for most antibiotic classes. Availability of a single reviewer to select, extract and critically appraise the studies.

**Conclusions:** This review provides the most up to date synthesis of evidence in relation to the risk of HCFA-CDI associated with exposure to specific antibiotic classes. Studies were predominantly conducted in North America or Europe and more studies outside of these settings are needed.

**Registration number:** Prospero CRD42020181817

## Introduction

*Clostridioides difficile* infection (CDI) is a leading cause of healthcare-associated infections and the most common cause of health care facility-associated (HCF) infectious diarrhoea in high-income countries. ^1^ *C. difficile* is a toxin-producing, anaerobic spore forming bacterium that is transmitted via the faecal-oral route. ^2^ Symptoms range from mild diarrhoea to life-threatening conditions such as pseudomembranous colitis and toxic megacolon, and recurrence occurs among approximately 20% of cases following an initial episode. ^2^ CDI is associated with a protracted length of stay incurring substantial direct and indirect healthcare utilisation costs. ^3–5^ Incidence rates of healthcare facility-associated (HCFA) CDI are geographically and temporally variable but have generally increased in the past 20 years. ^1, 6, 7^ Explanations for the increasing occurrence include increased testing and more sensitive diagnostic tests, increased use of broad-spectrum antibiotics, inadequate prevention measures, ageing population and emergence of community strains. ^7^

Antibiotic use is the most important modifiable risk factor for CDI. Antibiotic exposure alters the natural flora of the intestines allowing *C. difficile* to proliferate. Other important risk factors include advanced age, increased number of comorbidities or severe underlying disease, and duration of healthcare exposure. ^8, 9^ A longer duration of hospitalisation is correlated with advanced age and severity of underlying illness, and increases the probability of *C. difficile* acquisition from the environment and exposure to antibiotics. Although almost all antibiotics have been have been associated with CDI, clindamycin, cephalosporins, carbapenems and fluoroquinolones are most frequently associated with infection. ^10, 11^

Antibiotic stewardship reduces unnecessary antibiotic use in hospitals ^12^ and is crucial to controlling CDI in HCFs alongside surveillance, isolation precautions, hand hygiene and environmental cleaning. ^8, 13^ Limiting antibiotic use effectively reduces the incidence of CDI in HCFs, ^14^ however, CDI remains a significant problem and has been named by the US Centers for Disease Control as an urgent threat to public health. ^15^

It is therefore important to monitor changes in risk for HCFA-CDI associated with changes in antibiotic usage and changing *C.difficile* epidemiology. The objective of this systematic review is to update the evidence for epidemiological associations between specific antibiotic classes and HCFA-CDI. This review updates earlier reviews that covered studies conducted up to 2012, ^10, 11^ to include studies undertaken between 2013 and 2020.

## Methods

The study has been conducted in accordance with the PRISMA statement. ^16^ The objectives, inclusion criteria and methods for analysis were specified in advance and registered with PROSPERO (CRD42020181817).

### Eligibility criteria

Studies were eligible for inclusion in the review if they were conducted among hospital inpatients, measured any exposure to individual antibiotics or antibiotic classes prior to development of the outcome, included a comparison group, and measured the occurrence of HA-CDI as an outcome. Study designs eligible for inclusion consisted of case-control studies, cohort studies, analytical cross-sectional studies and randomised controlled trials (if they reported the risk associated with the exposure).

Studies were excluded if they investigated risk factors for severe disease, relapse or recurrence, asymptomatic colonisation, or were specific studies of paediatric populations. Studies that did not report exposure to specific antibiotics or antibiotic classes were also excluded, as were studies examining community associated infection or did not adequately exclude CA. Case reports, case series, and descriptive cross-sectional studies (i.e. those without comparison groups) were excluded. The review was limited to English-language publications, however, non-English articles were included in searches and their abstracts and full texts were assessed for eligibility.

### Information sources and searching

PubMed, Scopus and Web of Science Core Collection were searched on 21^st^ April 2020 for studies published since 1^st^ January 2013. WorldCat (www.worldcat.org) and Proquest Dissertations and Theses (PQDT www.proquest.com) were used to search for dissertations and theses. Final follow up searches to 31^st^ December 2020 were conducted on 11^th^ February 2021.

The search strategy used was consistent with the previous review. ^10^ The full search strategy for each database presented in the Supplementary file. Search results were exported to Endnote X9.1, which was used to identify and remove duplicates prior to importing to Covidence (www.covidence.org) for title and abstract screening.

### Study selection

Titles and abstracts were screened to eliminate irrelevant studies. Full text articles were then inspected for eligibility. Studies were classified as confirmed HCFA-CDI if they included a clear definition of HCFA-acquisition of CDI, or probable HCFA-CDI on the basis of information provided in the report, e.g. evidence that the minimum length of stay until onset of symptoms was >48 hours. Studies that had been excluded from the previous review because they did not use an explicit definition of HA were re-evaluated and subsequently included in this review if there was sufficient evidence that HA-CDI was probable.

### Data collection

Data were extracted from each study using a pro forma template that included the study characteristics (citation, country, setting, study period based on earliest time point, study population, study design, HCFA-CDI case and non-case definitions, antibiotic name, exposure period and timing, comparison group), number of subjects in exposure categories for cases and non-cases, effect estimates (odds ratio; relative risk) and 95% confidence intervals.

Antibiotic exposures were categorised into their main classes with additional sub-group categorisation for cephalosporins (1^st^, 2^nd^, 3^rd^, 4^th^ generation), penicillins (ampicillin-like drugs [aminopenicillins]; beta-lactamase inhibitor combinations; broad-spectrum [antipseudomonal] penicillins; penicillin G-like drugs [natural penicillins]) using the MSD Manual. ^17^ The unexposed comparator groups were categorised as no antibiotics; unexposed to antibiotic of interest; a reference antibiotic. The exposure period was classified in two ways. First, whether antibiotic administration before CDI diagnosis was recorded by the study and, second, the period of time over which antibiotic administration was measured and categorised as: during index admission; up to 1 month prior to CDI diagnosis; up to 1-2 months prior; up to 2-3 months prior; preoperative prophylaxis; during admission - prior to ICU; during ICU admission.

The non-CDI case groups were recorded as asymptomatic or symptomatic (patients had diarrhoea but tested negative for *C. difficile*). Study setting was classified according to geographic region (North America, Latin America, Europe, East Asia & Pacific) and patient population: general adult inpatients and specific clinical sub-groups (HA-pneumonia, HA-diarrhoea, antibiotic treatment, ICU patients, COPD patients, *C. difficile* colonised, haematology-oncology patients, surgical – gastrointestinal, surgical – non-gastrointestinal, type 2 diabetes).

### Assessment of methodological quality

The Newcastle–Ottawa Scale for the Assessment of Quality (NOS) was used to assess study quality. ^18^ The NOS comprises nine items across the domains of: selection, comparability, and exposure (case-control studies) or outcome (cohort studies). Each accomplished item receives one point and studies are classified as high quality (score 7–9), moderate quality (score 4–6), or poor quality (score 0–3). Important confounding domains considered in the study for assessment of comparability were identified from previous studies and included: age, comorbidities, severity of underlying disease (1 point), and healthcare facility exposure or other treatment (1 point) ^9^. Poor quality studies were not excluded from the review, instead overall study quality and level of comparability were explored as potential sources of heterogeneity.

### Analysis

Data were entered into Excel for coding and exported to Stata version 14 for analysis. A quantitative descriptive analysis summarising the characteristics of included studies was undertaken, presented as counts and percentages.

To assess the association between each antibiotic class and HA-CDI, a pooled random effects analysis using the DerSimonian-Laird inverse variance approach was used. ^19, 20^ The meta-analysis was restricted to studies that used a comparison group that was unexposed to the antibiotic of interest, as this was the comparison used in most studies. As most studies reported odds ratios (OR), the most fully adjusted odds ratio was used for synthesis. Where studies reported results for multiple antibiotics within each class, without providing the relevant overall association for that class, effect estimates were combined by taking the weighted average of the log ORs, with inverse variance weights. Heterogeneity of pooled effects was assessed using the I^2^ statistic, which describes the percentage of variability in the effect estimates that is due to heterogeneity rather than chance, and its chi-squared statistic for evidence of heterogeneity (p<0.10).

A sub-group analysis was used to assess study characteristics identified *a priori* based on previous findings as sources of heterogeneity. Specific characteristics investigated included setting (geographic region, time period when the study was conducted, patient population), exposure measurement (time period prior to CDI diagnosis, measurement after onset), and methodology (study design, HA definition, non-case comparison group, study quality, confounder adjustment). Only antibiotic classes with data from at least 10 studies were included in the sub-group analysis. The sub-group pooled associations and their corresponding I^2^ statistics and tau-squared statistics (for between study variance) were examined. The overall association between the study characteristic was investigated using random effects metaregression; variables associated with between study variation at p<=0.10 in a bivariate analysis were included in multivariable models.

Funnel plot analysis was used to assess bias due to missing results (publication bias). ^21^

## Results

The PRISMA diagram is presented in Figure 1. Of 804 titles and abstracts of non-duplicate articles screened, 197 full text articles were assessed for eligibility and 23 were included in the review along with 16 studies from the previous review covering studies until the end of 2012 (total 39). Of the 23 studies identified from the latest searches, only 11 studies reported a clear definition of hospital acquisition with a further 12 deemed probable HCFA-CDI based on information available in the article. Two papers previously excluded from the earlier review (consisting of 14 studies) were identified as probable HCFA-CDI and subsequently added to this update.

**Figure 1.**
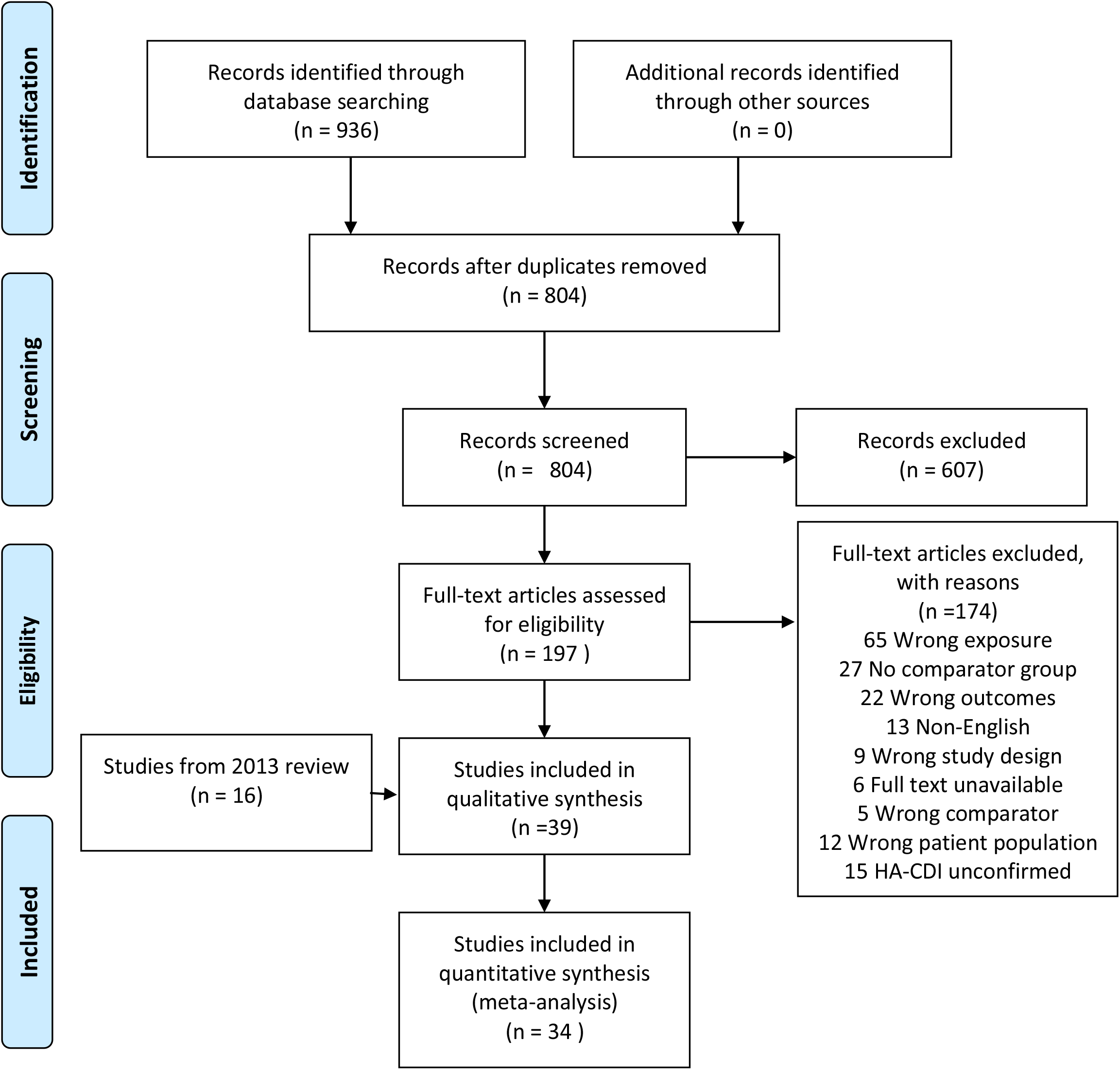
PRISMA flow diagram of study selection.

Four studies of patients undergoing solid organ transplants ^22–25^ and five stem cell transplant studies ^26–30^ were excluded because of the long follow times after discharge precluding establishment of HA. For example, in the study by Cusini et al. ^22^ kidney transplant patients were followed for 2 years of which only 56% of CDI cases were deemed nosocomial.

### Description of studies

The characteristics of the included studies identified are summarised in Table 1. Full details of each study identified are provided in the supplementary file (Table S1). Most studies were either cohort or case-control studies, where cohort studies were the predominant design in articles published since 2013, compared to case-control studies in earlier articles. Compared to the earlier review, a more diverse range of patient groups was investigated in the recent studies and from a wider range of geographic regions. Of the 18 cohort studies, 16 were clinical cohort studies, examining specific inpatient sub-groups. Most studies at both time points used patients not diagnosed with CDI as the non-case group, with 23% using symptomatic non-cases. Most studies (87%) used non-exposure to the antibiotic of interest as the unexposed comparator.

**Table 1.** Characteristics of studies examining associations between antibiotics and HA-CDI included in the 2020 systematic review update.

|  | Number of studies (%) |  |  |
| --- | --- | --- | --- |
|  | 2013 review* | 2020 update | Total |
| Number of studies | 16 | 23 | 39 |
| Region |  |  |  |
| North America | 11 (68.7) | 9 (39.1) | 20 (51.3) |
| Latin America | 0 (0.00) | 3 (13.0) | 3 (7.7) |
| Europe | 4 (25.0) | 7 (30.4) | 11 (28.2) |
| East Asia & Pacific | 1 (6.2) | 4 (17.4) | 5 (12.8) |
| Study population |  |  |  |
| All inpatients | 11 (68.7) | 6 (26.1) | 17 (43.6) |
| HA-pneumonia | 1 (6.2) | 2 (8.7) | 3 (7.7) |
| HA-diarrhoea | 0 (0.0) | 2 (8.7) | 2 (5.1) |
| Antibiotic treated | 4 (25.0) | 4 (17.4) | 8 (20.5) |
| ICU patients | 0 (0.0) | 2 (8.7) | 2 (5.1) |
| COPD inpatients | 0 (0.0) | 1 (4.3) | 1 (2.6) |
| *C.difficile* colonisation | 0 (0.0) | 1 (4.3) | 1 (2.6) |
| Haematology-oncology patients | 0 (0.0) | 1 (4.3) | 1 (2.6) |
| Surgical | 0 (0.0) | 3 (13.0) | 3 (7.7) |
| Type 2 diabetes inpatients | 0 (0.0) | 1 (4.3) | 1 (2.6) |
| Outbreak investigation |  |  |  |
| No | 13 (81.2) | 22 (95.6) | 35 (89.7) |
| Yes | 3 (18.7) | 1 (4.3) | 4 (10.3) |
| Study design |  |  |  |
| Cohort | 2 (12.5) | 16 (69.6) | 18 (46.1) |
| Case-control | 13 (81.2) | 5 (21.7) | 18 (46.1) |
| Nested case-control | 1 (6.2) | 0 (0.0) | 1 (2.6) |
| Case cohort | 0 (0.0) | 2 (8.7) | 2 (5.1) |
| Non-case group* |  |  |  |
| Symptomatic | 2 (12.5) | 7 (30.4) | 9 (23.1) |
| Non-CDI | 13 (81.2) | 14 (60.9) | 27 (69.2) |
| Two control groups | 1 (6.2) | 2 (8.7) | 3 (7.7) |
| Unexposed group |  |  |  |
| No antibiotics | 0 (0.0) | 1 (4.3) | 1 (2.6) |
| Unexposed to specific antibiotic | 16 (100) | 18 (78.3) | 34 (87.2) |
| Reference antibiotic | 0 (0.0) | 4 (17.4) | 4 (10.3) |
| Quality |  |  |  |
| Low (0-3) | 0 (0.0) | 1 (4.3) | 1 (2.6) |
| Moderate (4-6) | 3 (18.7) | 10 (43.5) | 13 (33.3) |
| High (7-9) | 13 (81.2) | 12 (52.2) | 25 (64.1) |

Abbreviations: HA hospital-acquired; COPD chronic obstructive pulmonary disease
\*Includes two studies identified but not included in the 2013 review.
\*Symptomatic refers to patients with diarrhoea who tested negative for *C.difficile*. Non-CDI refers to all patients that were not diagnosed with CDI. May include patients without diarrhoea and also patients with diarrhoea plus a negative *C.difficile* test.

### Quality appraisal

Most studies were graded as medium or high quality (Table 1). Full details of the NOS quality appraisal score for each study is summarised in the supplementary file (Table S2). Selection and confounding were important sources of error, with only 17 (43%) studies attaining the maximum score for selection and 15 (38%) studies reaching the maximum score for comparability; 86% of case-control designs scored 3/3 for exposure measurement, and 50% of cohort designs scored 3/3 for outcome measurement.

### Study results and pooled effects

Studies that used an exposure comparison group consisting of those not exposed to the antibiotic measured were included in meta-analyses (n=34).

For non-beta-lactam antibiotic classes (Figures 2 and 3), the strongest evidence for an association was seen for quinolones (fluoroquinolones) and lincosamides (clindamycin). Overall, quinolones were associated with a 34% increased odds of HCFA-CDI (OR=1.34, 95% CI=1.13-1.60), although individual ORs ranged from 0.15 to 15.30. Excluding the study with an extremely small outlier association (0.15) made little change to the pooled result (OR=1.37, 95%CI=1.15-1.63; I^2^=85.4%). Lincosamides were associated with a 56% increased odds of HCFA-CDI (OR=1.56, 95% CI=1.13-2.14), with individual study ORs ranging from 0.39 to 9.10. Weak positive pooled associations were observed for aminoglycosides, macrolides, and sulphonamides-trimethoprim, and a wide range in individual study effect sizes was found. There was strong heterogeneity observed for each of these classes, with ORs distributed in both positive and negative directions. Associations were found also for antibiotics used to treat CDI, particularly vancomycin (class glycopeptides) with a pooled OR of 1.91 (95%CI 1.32-2.78).

**Figure 2.**
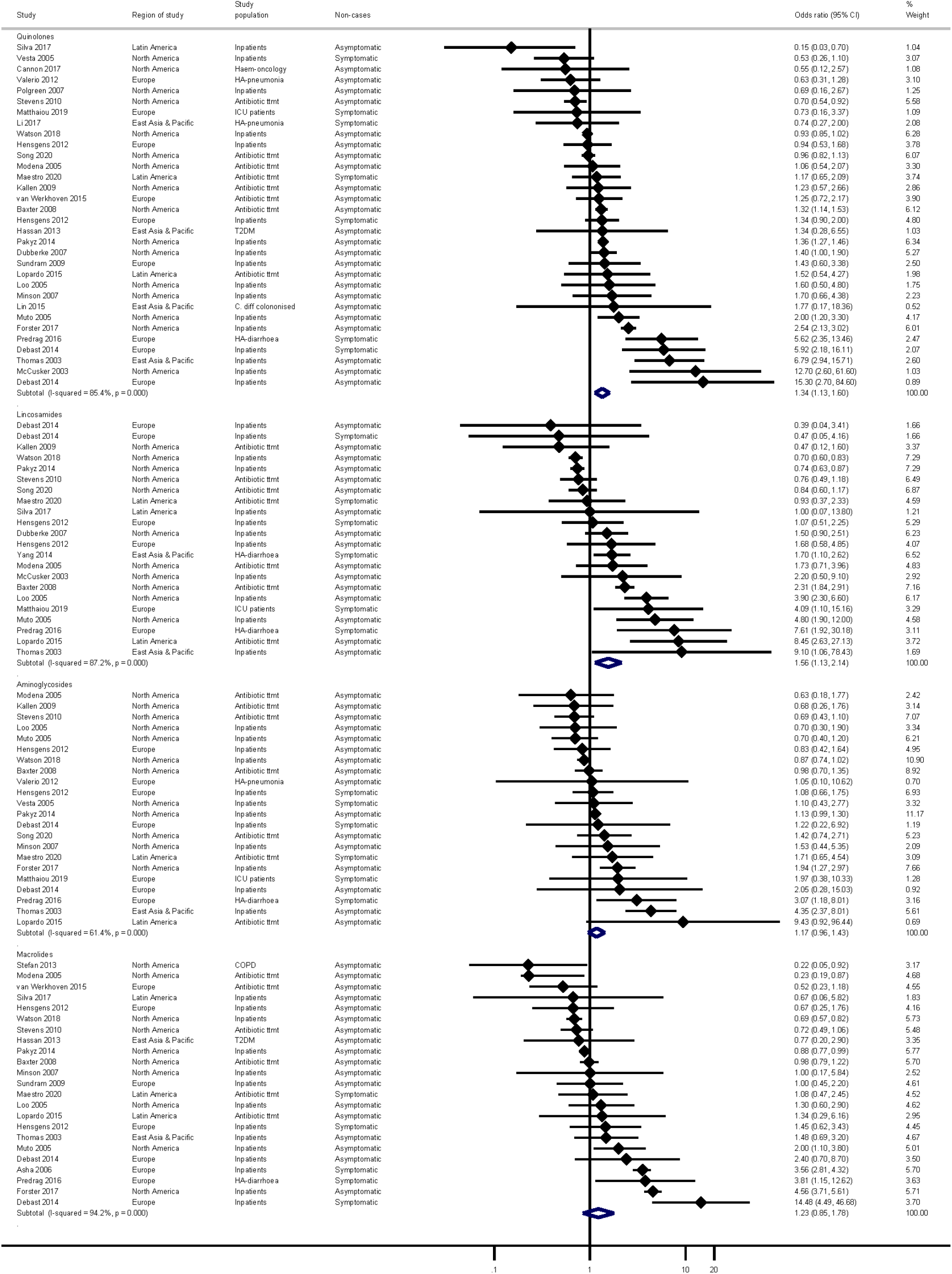
Forest plot for non beta lactam antibiotic classes: quinolones, aminoglycosides, macrolides, lincosamides.

**Figure 3.**
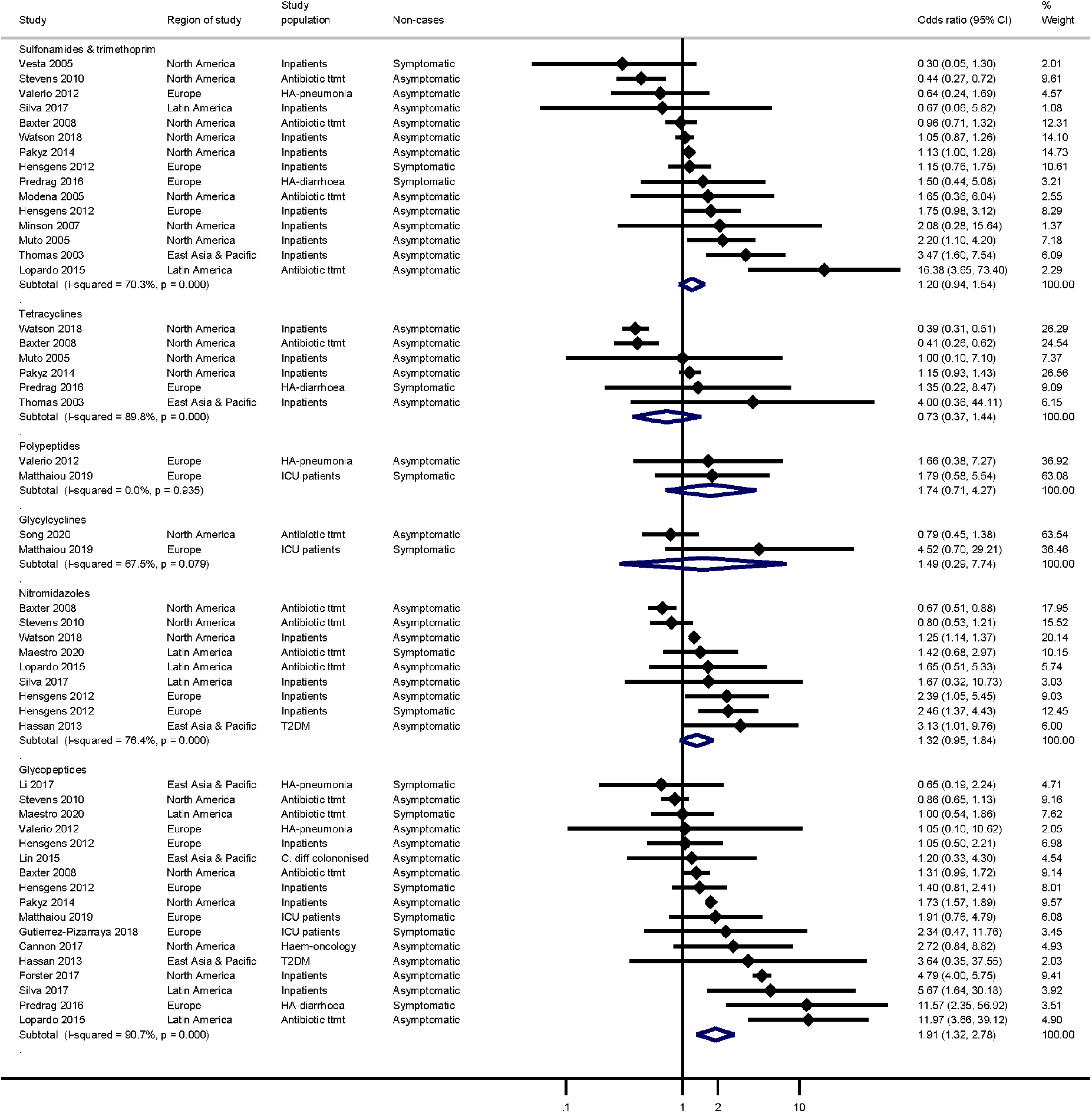
Forest plots for non beta lactam antibiotic classes: tetracyclines, sulfamethoxaxole and trimethoprim, miscellaneous.

Figure 4 displays the results for beta-lactam classes. The strongest association was seen for carbapenems (OR=2.55, 95%CI=1.83-3.55), with penicillins and cephalosporins associated with 33% and 79% increased odds of HA-CDI, respectively. There was substantial heterogeneity for all three main classes. For carbapenems, all ORs except one were in the positive direction ranging from 0.79 to 14.13. For penicillins and cephalosporins there were a number of ORs in opposite directions.

**Figure 4.**
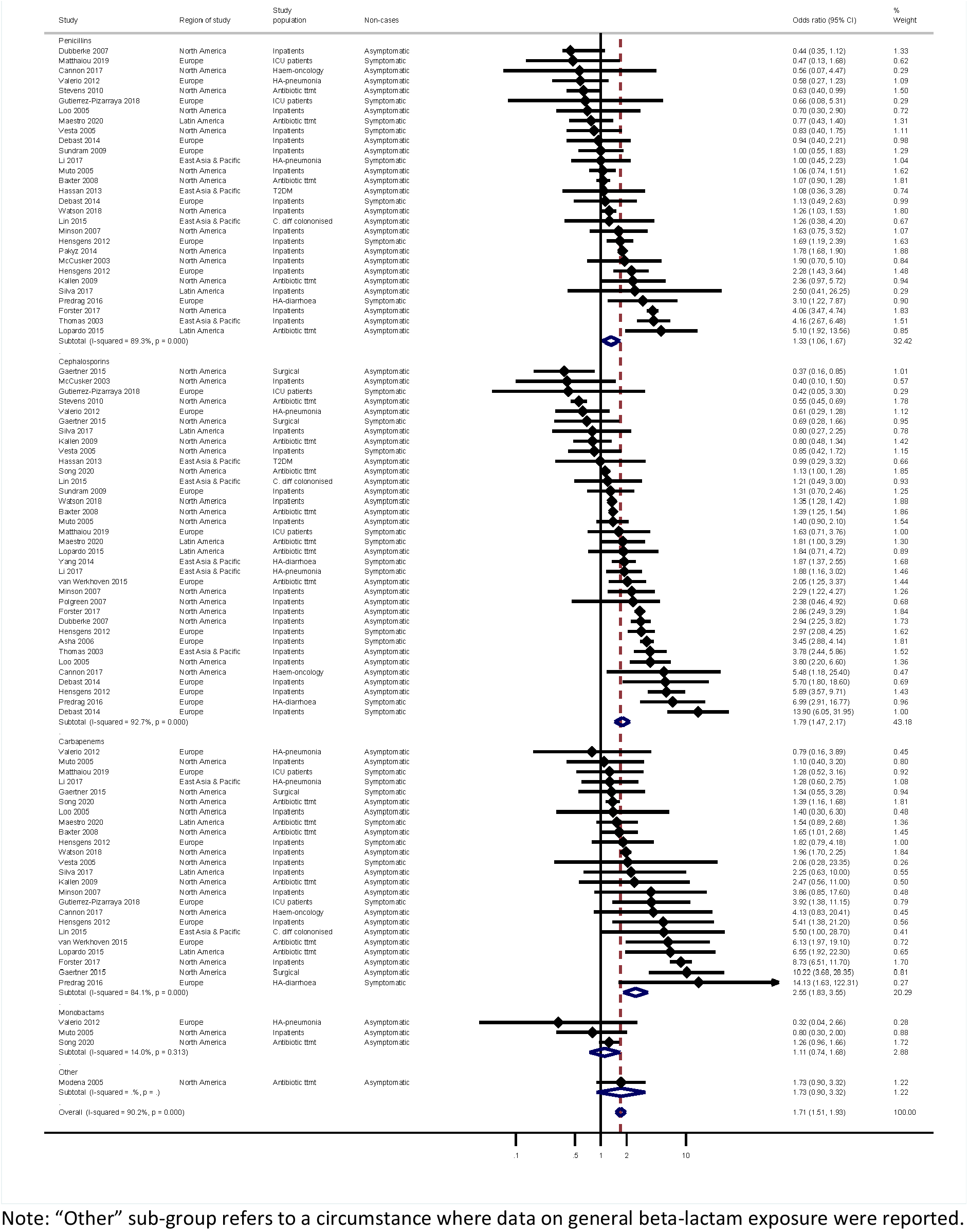
Forest plots for beta-lactam antibiotic classes.

Beta-lactamase inhibitor combination penicillin antibiotics were the most frequently reported penicillin sub-class (Figure 5). ORs ranged from 0.47 to 17.40 (three studies had OR<1) with a pooled OR of 1.43 (95%CI 1.16-1.77). There was little evidence of an association for aminopenicillins, and few studies reported data for broad-spectrum or penicillinase-resistant penicillins.

**Figure 5.**
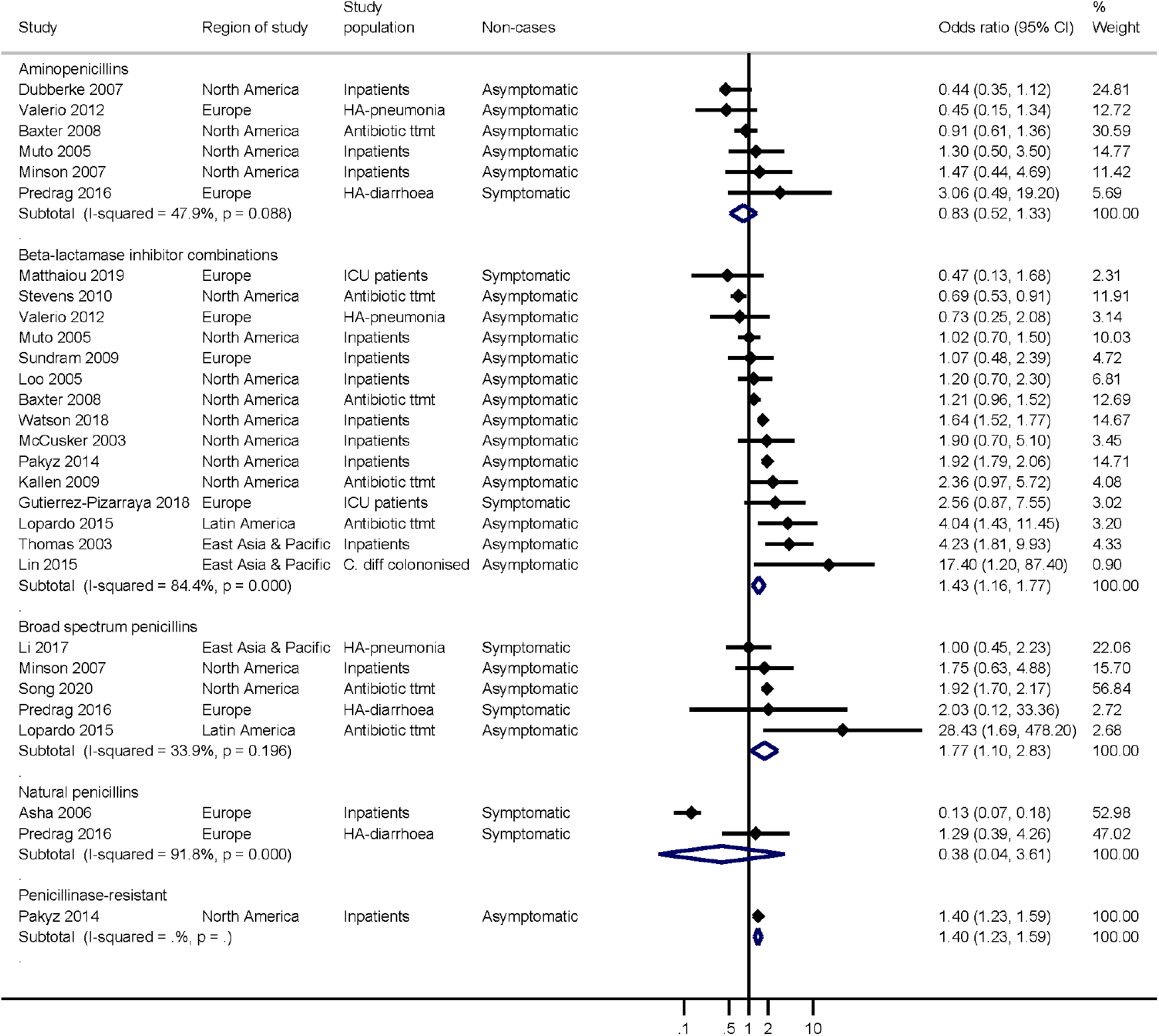
Forest plots for penicillin sub-classes.

For cephalosporins, 3^rd^ and 4^th^ generation classes were associated with a doubling of the odds of HA-CDI, 2^nd^ generation cephalosporins with a 58% increased odds of HA-CDI, and no evidence for an association with 1^st^ generation cephalosporins (Figure 6). The individual study findings for 4^th^ generation cephalosporins were the most homogenous, with all effect sizes in a positive direction and ranging from 1.1 to 3.24.

**Figure 6.**
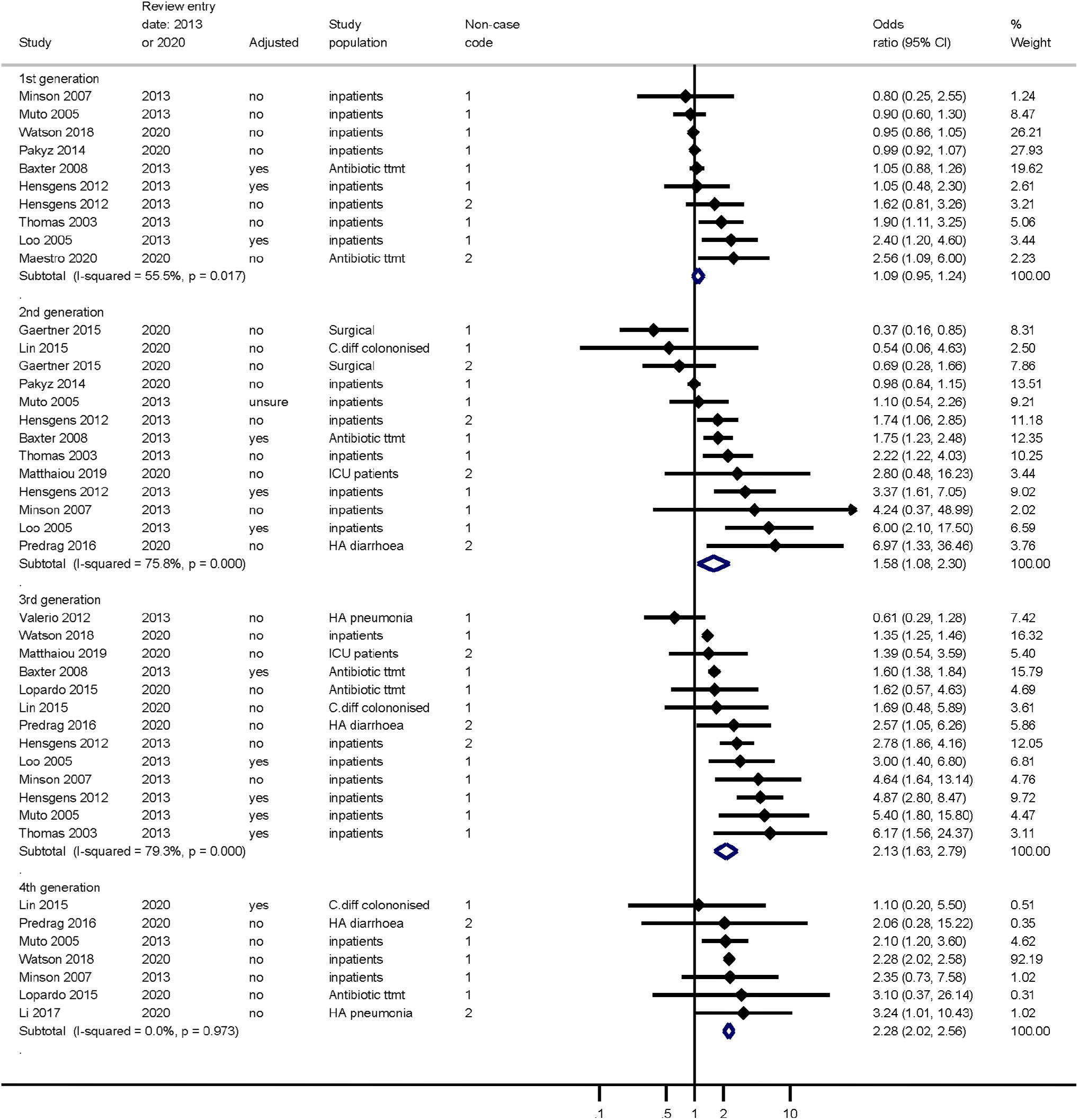
Forest plots for cephalosporin sub-classes.

### Metaregression and sub-group analyses

Ten antibiotic classes with a minimum of 10 studies and where more than 50% of the variation was due to heterogeneity were included in the sub-group analysis and metaregression. The results of subgroup analyses are presented as supplementary figures (Figures S1-S3). Although several sources of heterogeneity were identified for eight antibiotic classes, there were no sources common to all classes (Table 2). The most common sources were geographic region, exposure measurement period, and measurement of exposure after onset of symptoms.

**Table 2.**
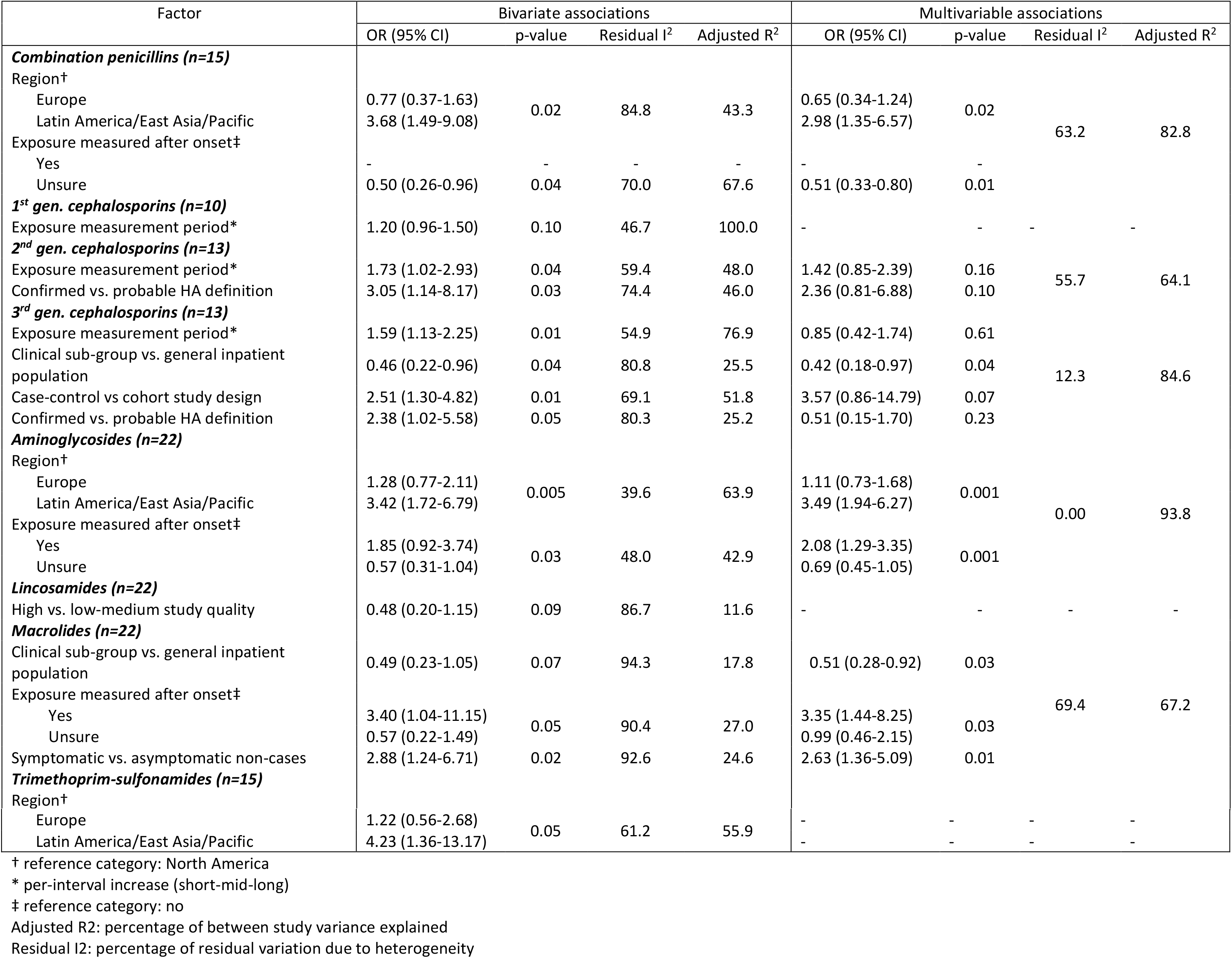
Metaregression analyses.

Studies conducted in Latin America, East Asia and Pacific regions reported stronger associations for combination penicillins, aminoglycosides and trimethoprim-sulfonamides, approximately 3-4 times higher than those reported by North American studies. Measurement of the antibiotic exposure after onset of symptoms was also a source of heterogeneity for aminoglycosides, with associations twice that of studies that collected exposure information preceding onset of symptoms. A longer window of antibiotic exposure measurement was associated with all three cephalosporin sub-classes in unadjusted analyses, and associations were attenuated in multivariable models. Other sources of heterogeneity for cephalosporins included definition of HCF acquisition (2^nd^ and 3^rd^ generation), study population (3^rd^ generation) and study design (3^rd^ generation). Studies using patient sub-groups tended to have weaker associations for a number of antibiotic classes, and this was a strong source of heterogeneity for associations with macrolides and 3rd generation cephalosporins, with effect sizes half of that seen in studies of general inpatients.

### Other findings

Five studies were not included in the meta-analysis due to differences in antibiotic exposure measurement; two Canadian studies used no antibiotic exposure as the reference category, ^31, 32^ one study examined duration and dosage of antibiotic exposure ^33^ and two UK studies used historical controls to examine the impact of changes to antibiotic formulary changes. ^34, 35^ The main results are summarised below.

A case-cohort study of adult inpatients of a large tertiary hospital in Canada measured the risk of HA-CDI associated with exposure to various antibiotic classes compared to patients with no antibiotic exposure in the five days prior to developing symptoms. ^31^ After adjusting for age, gender, hospital exposure, and infection pressure, the incidence of CDI was 2-3 times higher for penicillins, cephalosporins, fluoroquinolones, and carbapenems. The second Canadian study compared the risk of CDI for various antibiotics according to whether CDI developed during or following cessation of exposure compared to no exposure, taking into account competing events of death or discharge. ^32^ They found the risk of CDI was greatest in the period following exposure to penicillins, quinolones, macrolides, aminoglycosides or clindamycin.

The cohort study of ICU patients with hospital-acquired pneumonia by Li *et al*. ^33^ reported the mean duration of treatment and total dose per patient to be higher in patients who developed CDI for broad-spectrum cephalosporins, carbapenem, and oxacephems, although not all of the differences were statistically significant.

Two UK clinical cohort studies used historical controls to examine the impact of changes to antibiotic formulary changes. In patients with severe hospital-acquired pneumonia, none of the patients treated with amoxicillin plus temocillin (n=98) developed CDI compared to 7.4% (7/94) of patients who had previously been treated with piperacillin-tazobactam (both beta-lactamase inhibitor combinations). ^34^ In a study comparing gentamicin (aminoglycoside) with cefuroxime (2^nd^ generation cephalosporin) for total hip and knee replacement surgery prophylaxis, there were no CDI cases in the gentamicin group (n=2101) compared to 11 cases (0.2%) in the cefuroxime group (n=6094). ^35^

### Non-English language studies

Thirteen non-English language articles were identified in the current review of which only one study was eligible for inclusion in the review. ^36^ The case-control study of immunosuppressed patients reported exposure to fluoroquinolones was higher in CDI cases (36%) than controls (28%) and exposures to 2^nd^ or 3^rd^ generation cephalosporins was lower in cases (6%) than controls (14%). Details of the non-English language studies are summarised in Table S3.

### Funnel plot symmetry

Supplementary Figure S4 displays the funnel plots for the studies included in the meta-analyses of each main antibiotic class. Although there was not strong evidence for bias due to non-reporting of small studies, several of the plots reflect asymmetry associated with the heterogeneity previously addressed.

## Discussion

This systematic review updates the findings from two previous reviews through to 2020. Twenty-one studies published since 2013 were included in this review, in addition to 16 studies identified in earlier reviews. ^10, 11^ The main findings indicate that carbapenems and 3^rd^ and 4^th^ generation cephalosporin antibiotics remain most strongly associated with HCFA-CDI, with cases more than twice as likely to have recent exposure to these antibiotics prior to developing CDI. Modest associations (OR>1.5) were observed for quinolones (predominantly fluoroquinolones), lincosamides (namely clindamycin), 2^nd^ generation cephalosporins, and beta-lactamase inhibitor combination penicillin antibiotics. Individual study effect sizes were variable and heterogeneity was a common problem observed for most antibiotic classes. However, the risk of CDI for a given antibiotic will depend on the local prevalence of strains that are resistant to the particular antibiotic and a certain degree of heterogeneity is therefore to be expected. ^8^

Since 2013, there have been more studies conducted in a wider range of countries, including three from Latin America (covering Argentina, Mexico, Brazil) ^37, 38^ and four from East Asia and Pacific countries (China, Korea, Malaysia, Taiwan). ^33, 39–41^ The studies from Latin America and East Asia-Pacific regions reported stronger associations for a number of antibiotic classes with HCFA-CDI. With only a small number of studies from these regions conclusions regarding such differences are limited and further studies are needed.

In addition, a more diverse range of patient populations have been studied since 2013, with an increase in focused studies on clinical patient groups, such as surgical ^35, 42, 43^ and ICU patients, ^44^ or patients with particular conditions such as HA-pneumonia, ^33, 34, 45^ compared to earlier studies that predominantly studied general inpatients. Studies using patient sub-groups tended to have weaker associations. A potential explanation is that studies of patient sub-groups had shorter periods of antibiotic exposure measurement. For example, in studies assessing 3^rd^ generation cephalosporins, three out of seven studies of general inpatients measured exposure up to three months prior to CDI, whereas no study that was restricted to patient sub-groups used a three-month exposure window. Studies that used longer windows of exposure measurement had stronger effect sizes which could reflect a protracted time at risk for CDI following antibiotic treatment. Previous studies have shown that the risk of CDI declines between 1-2 months following cessation of antibiotic treatment. ^31, 46^ Alternatively, it could mean that these studies simply picked up more exposed individuals, resulting in differential misclassification if this was not consistent for cases and non-cases.

Another exposure measurement source of heterogeneity was the inadequate description or failure to limit recording of antibiotic exposure to a period preceding onset or diagnosis of CDI, producing biased effect estimates. This could explain the associations found for antibiotics used to treat CDI such as vancomycin and metronidazole (glycopeptide and nitromidazole classes, respectively), although this could not be established in this review, and both are able to incite CDI. ^47^ Future studies should clearly report the parameters of exposure measurement, including all sources of information on exposure, as it was often unclear particularly in studies with a longer exposure window whether in-hospital prescription only was recorded, or whether prescription in the community setting was included. Information on dose-response relationships is generally lacking and studies investigating the risk of HCF-CDI associated with the timing and duration of antibiotic exposure are needed.

Other methodological differences in studies, such as the use of a clear definition of hospital-acquisition, the choice of non-case comparison group, and group comparability on confounding factors, were not consistent sources of heterogeneity, but had varying influences on study results. A variety of diagnostic laboratory testing methods that have varying sensitivity and specificity were used by studies included in the review. With increasing computing power, cohort studies utilising routinely collected patient data have increased in recent years - investigators should clearly report surveillance definitions, criteria and laboratory methods for diagnosis of CDI in their institutions, and how these align with relevant current guidelines. ^8, 48^ Studies are subject to diagnostic suspicion bias if patients exposed to antibiotics are more likely to be tested for *C. difficile*.

This review provides the most up to date synthesis of evidence in relation to the risk of HA-CDI associated with exposure to specific antibiotic classes. However, the review is limited by the availability of a single reviewer to select, extract and critically appraise the studies. Since the previous review that covered studies published up to the end of 2012, there has been a marked increase in the overall number of studies eligible for inclusion. As found previously, studies were variable in methodological quality and quality of reporting results. Studies from the Latin America and East Asia-Pacific regions have increased, although studies were predominantly conducted in North America or Europe, and more studies outside of these settings are needed.

## Data Availability

Request from author.

---

**Literature search, performed 21st April 2020-total retrieved 893**

| No. | Database | Query | Results |
| --- | --- | --- | --- |
| 1 | Pubmed | (clostridium difficile AND (diarrhea OR colitis) AND (anti-infective agents OR antimicrobial agents OR antibiotic) AND (epidemiologic studies) NOT review[pt] AND (humans[Filter])) AND (("2013/01/01"[Date - Publication] : "2020/04/21"[Date - Publication]) AND (humans[Filter]))<br>Translation:<br>(((((("clostridium difficile"[MeSH Terms] OR ("clostridium"[All Fields] AND "difficile"[All Fields])) OR "clostridium difficile"[All Fields]) AND (((("diarrhea"[MeSH Terms] OR "diarrhea"[All Fields]) OR "diarrheas"[All Fields]) OR "diarrhoea"[All Fields]) OR "diarrhoeas"[All Fields]) OR ((("colitis"[MeSH Terms] OR "colitis"[All Fields]) OR "colitides"[All Fields])))) AND ((((((("anti infective agents"[Pharmacological Action] OR "anti-infective agents"[MeSH Terms]) OR ("anti infective"[All Fields] AND "agents"[All Fields])) OR "anti infective agents"[All Fields]) OR ((("anti"[All Fields] AND "infective"[All Fields]) AND "agents"[All Fields])) OR "anti infective agents"[All Fields]) OR (((("anti infective agents"[Pharmacological Action] OR "anti-infective agents"[MeSH Terms]) OR ("anti infective"[All Fields] AND "agents"[All Fields])) OR "anti infective agents"[All Fields]) OR ("antimicrobial"[All Fields] AND "agents"[All Fields])) OR "antimicrobial agents"[All Fields])) OR ((((((("anti bacterial agents"[Pharmacological Action] OR "anti-bacterial agents"[MeSH Terms]) OR ("anti bacterial"[All Fields] AND "agents"[All Fields])) OR "anti bacterial agents"[All Fields]) OR "antibiotic"[All Fields]) OR "antibiotics"[All Fields]) OR "antibiotic s"[All Fields]) OR "antibiotical"[All Fields])))) AND ((("epidemiologic studies"[MeSH Terms] OR ("epidemiologic"[All Fields] AND "studies"[All Fields])) OR "epidemiologic studies"[All Fields])) NOT "review"[Publication Type]) AND "humans"[MeSH Terms]) AND (2013/1/1:2020/4/21[Date - Publication] AND "humans"[MeSH Terms]) | 339 |
| 2 | Scopus | (TITLE-ABS-KEY("clostridium difficile" OR "clostridioides difficile") AND TITLE-ABS-KEY(diarrhe?a OR colitis) AND TITLE-ABS-KEY(anti-infective OR antimicrobial OR antibiotic) AND TITLE-ABS-KEY("case?control" OR cohort OR epidemiology)) AND PUBYEAR > 2012 | 181 |
| 3 | Web of Science Core Collection | TS=(((("clostridium difficile" OR "clostridioides difficile") AND ( "diarrhe?a" OR colitis) AND ("anti-infective" OR antibiotic OR antimicrobial) AND ( "case?control" OR cohort OR retrospective OR prospective))))<br>Refined by: [excluding] DOCUMENT TYPES: ( REVIEW OR EDITORIAL MATERIAL )<br>*Indexes=SCI-EXPANDED, SSCI, A&HCI, CPCI-S, CPCI-SSH, ESCI, CCR-EXPANDED, IC Timespan=2013-2020* | 134 |
| 4 | WorldCat | 'kw:("clostridium difficile" OR "clostridioides difficile") AND ("public health" OR "medicine" OR "epidemiology")' > '2013..2020' > 'Thesis/dissertation' | 82 |
| 5 | Proquest Dissertations and Theses | "clostridium difficile" AND su.Exact("epidemiology") AND (antibiotic OR antimicrobial)<br>*Date range: 01/01/2013 to to 21/04/2020* | 157 |

**Literature search updates to 31^st^ December 2020 (performed 11/02/2021) – total retrieved 43**

| No. | Database | Query | Results |
| --- | --- | --- | --- |
| 1 | Pubmed | (clostridium difficile AND (diarrhea OR colitis) AND (anti-infective agents OR antimicrobial agents OR antibiotic) AND (epidemiologic studies) NOT review[pt] AND (humans[Filter])) AND (("2020/04/21"[Date - Publication] : "2020/12/31"[Date - Publication]) AND (humans[Filter])) | 12 |
| 2 | Scopus | (TITLE-ABS-KEY("clostridium difficile" OR "clostridioides difficile") AND TITLE-ABS-KEY(diarrhe?a OR colitis) AND TITLE-ABS-KEY(anti-infective OR antimicrobial OR antibiotic) AND TITLE-ABS-KEY("case?control" OR cohort OR epidemiology)) AND PUBDATETXT("April 2020" or "May 2020" or "June 2020" or "July 2020" or "August 2020" or "September 2020" or "October 2020" or "November 2020" or "December 2020") | 9 |
| 3 | Web of Science Core Collection | TS((((("clostridium difficile" OR "clostridioides difficile") AND ( "diarrhe?a" OR colitis) AND ("anti-infective" OR antibiotic OR antimicrobial) AND ( "case?control" OR cohort OR retrospective OR prospective) ))))<br>Indexes=SCI-EXPANDED, SSCI, A&HCI, CPCI-S, CPCI-SSH, ESCI, CCR-EXPANDED, IC Timespan=2020 | 18 |
| 4 | WorldCat | kw:("clostridium difficile" OR "clostridioides difficile") AND ("public health" OR "medicine" OR "epidemiology")' > '2020..2020' > 'Thesis/dissertation' | 4 |
| 5 | Proquest Dissertations and Theses | "clostridium difficile" AND su.Exact("epidemiology") AND (antibiotic OR antimicrobial)<br>*Additional limits: 21/04/2020 to 31/12/2020* | 0 |

**Table S1.**
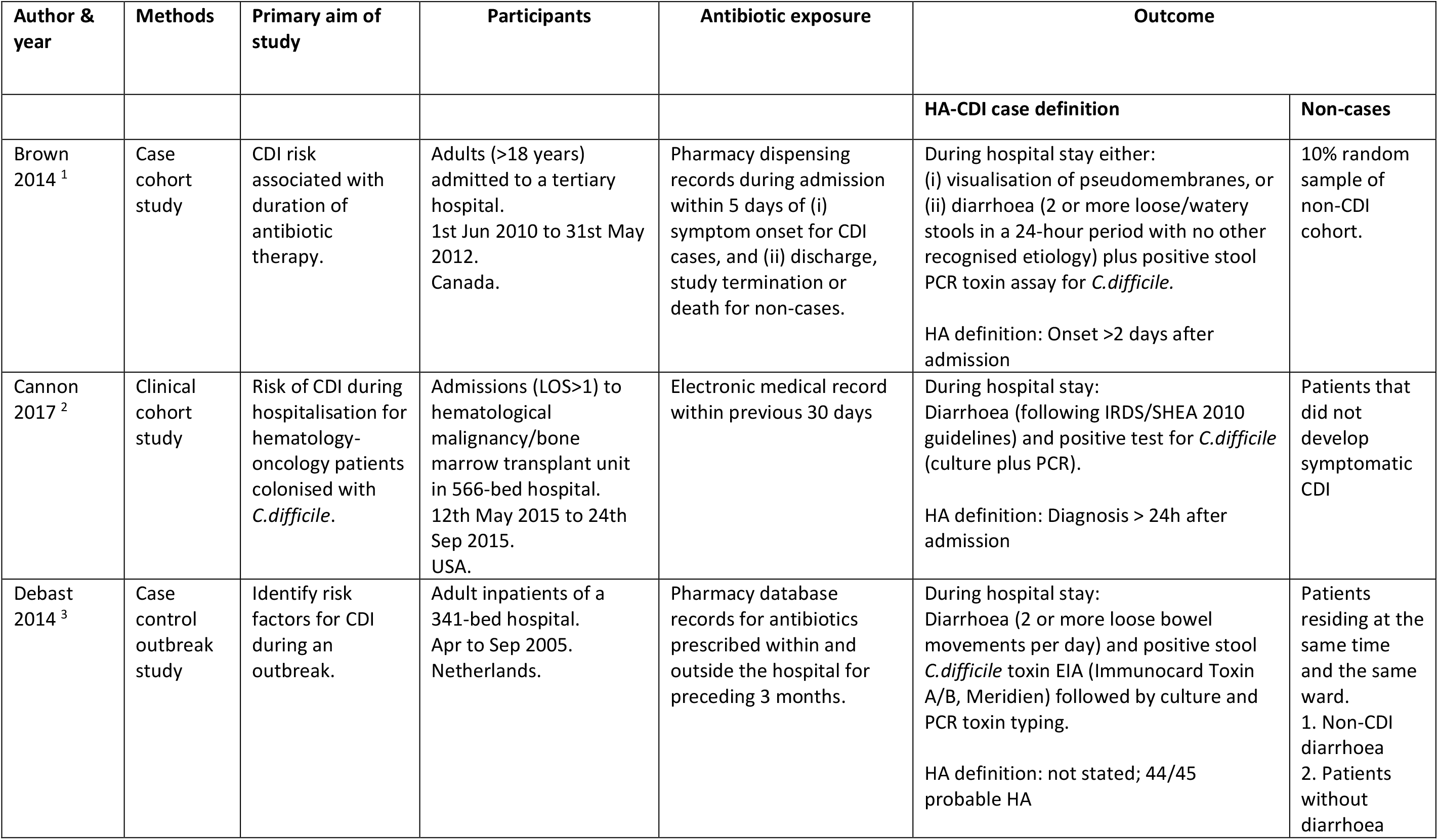
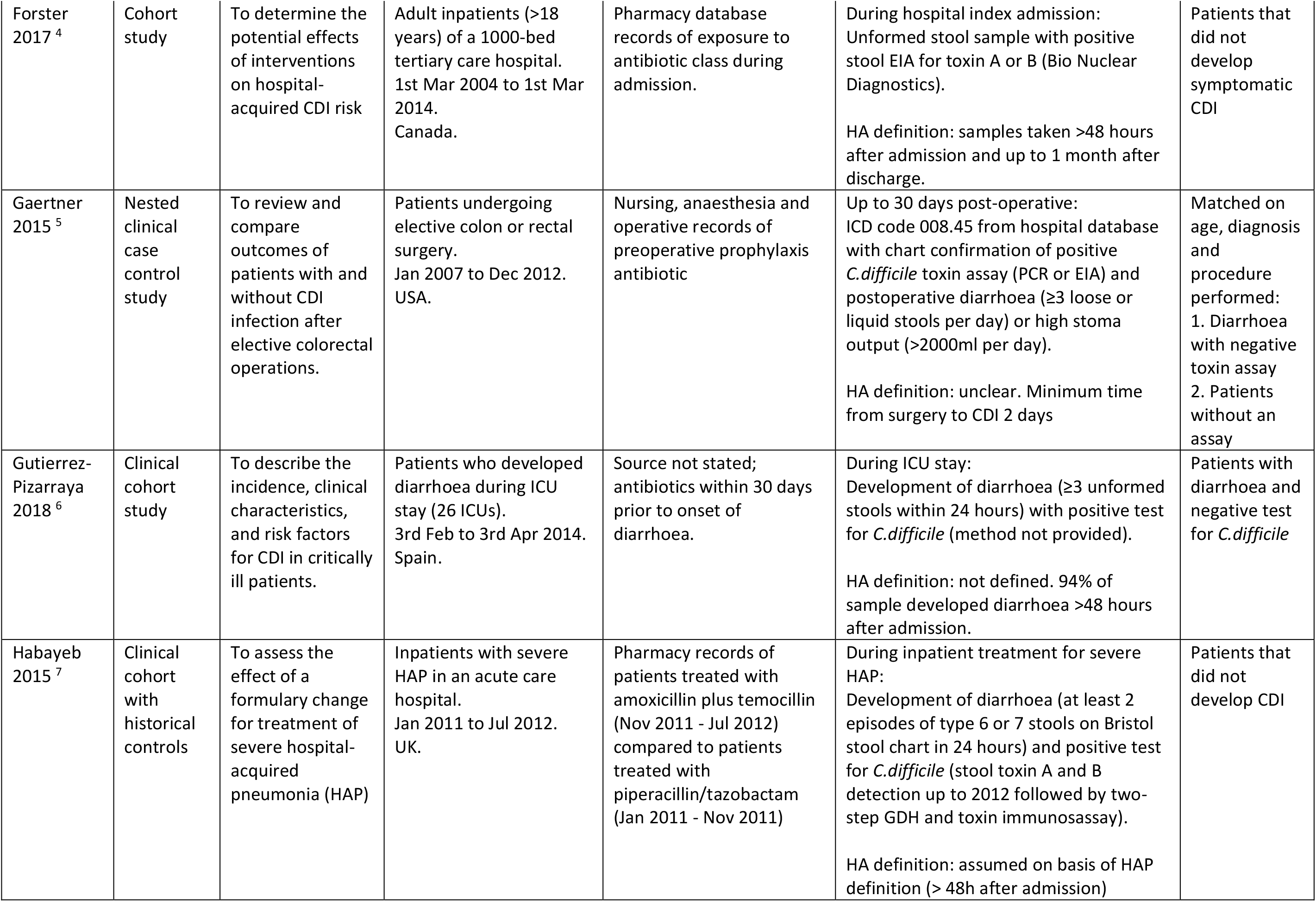
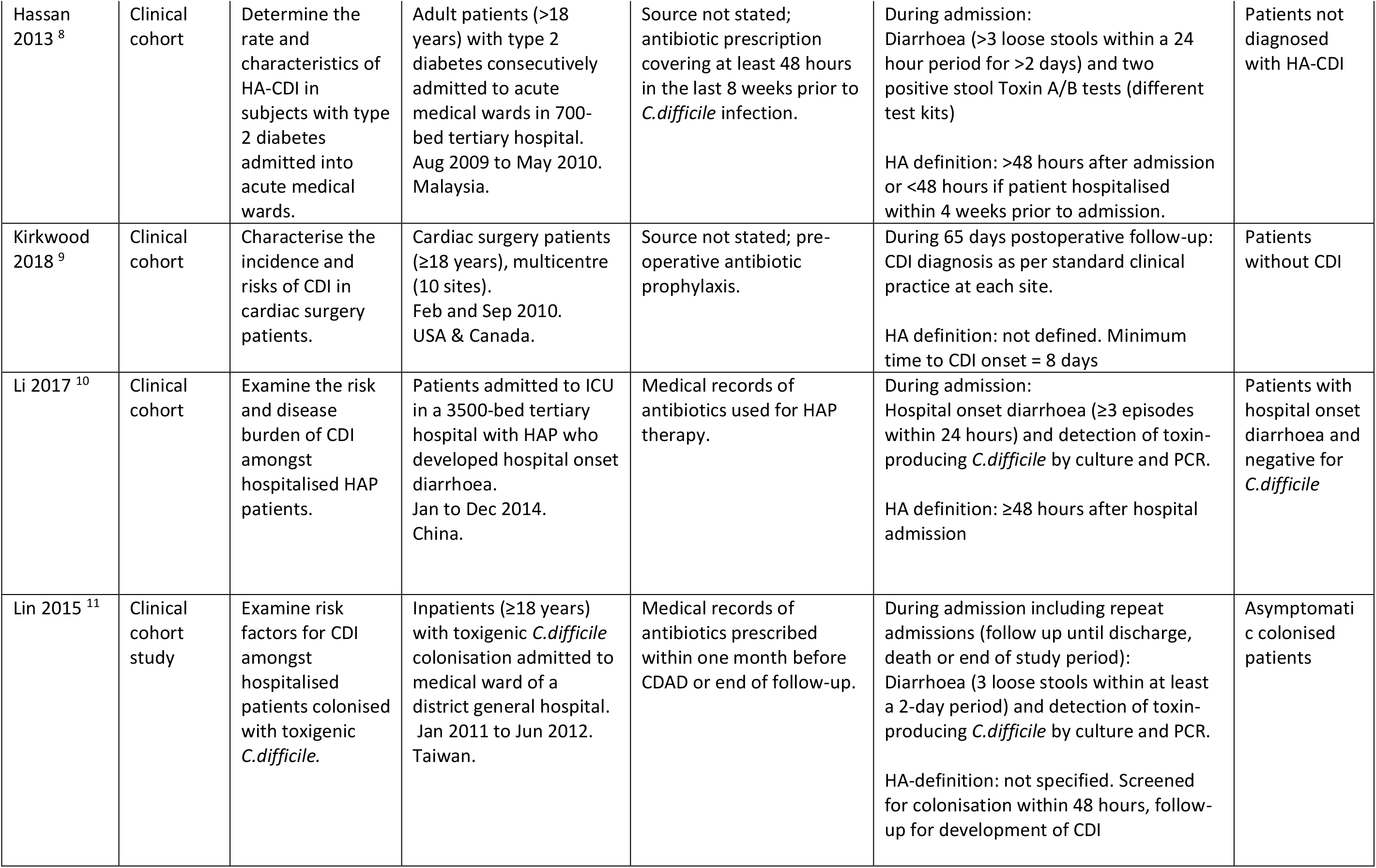
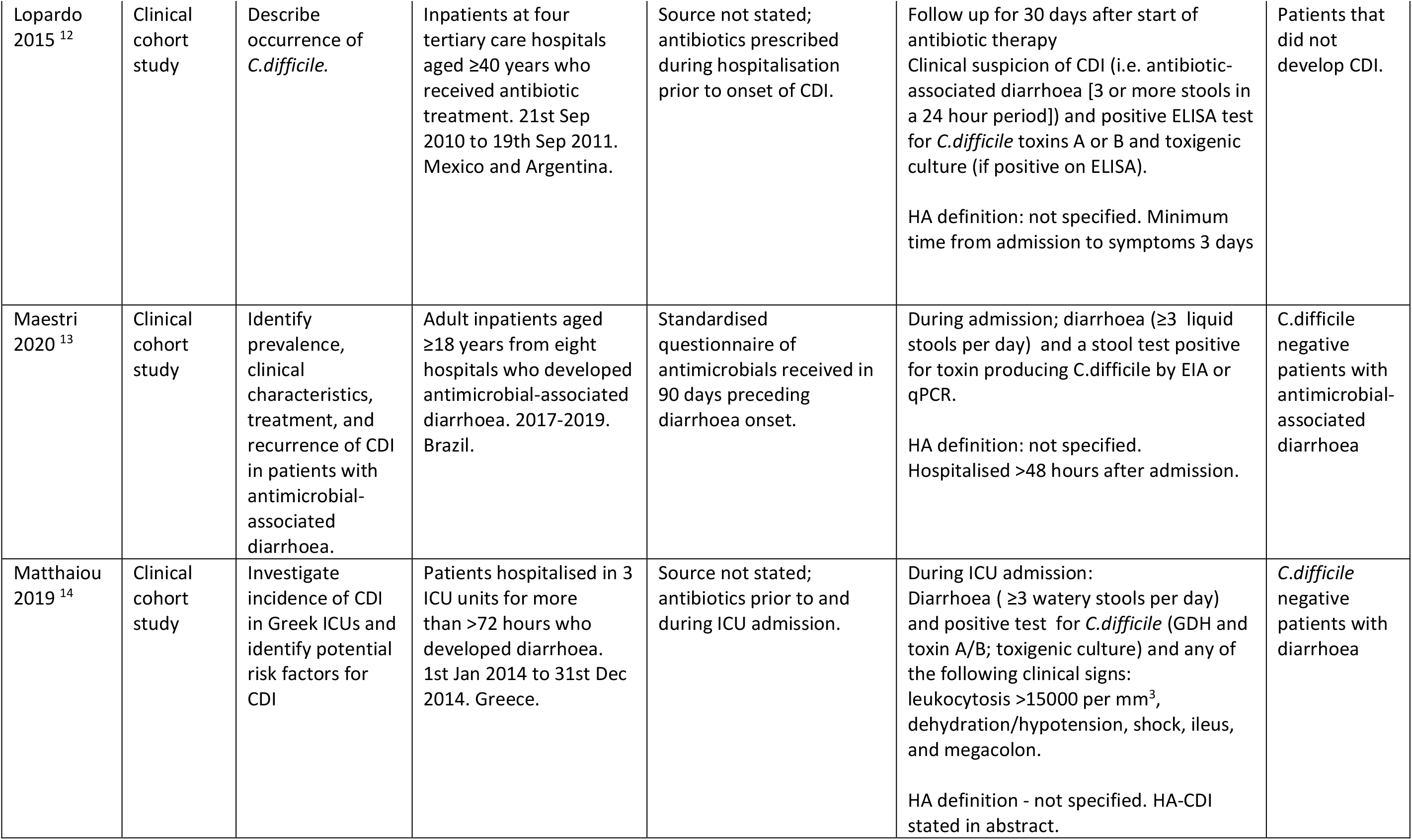
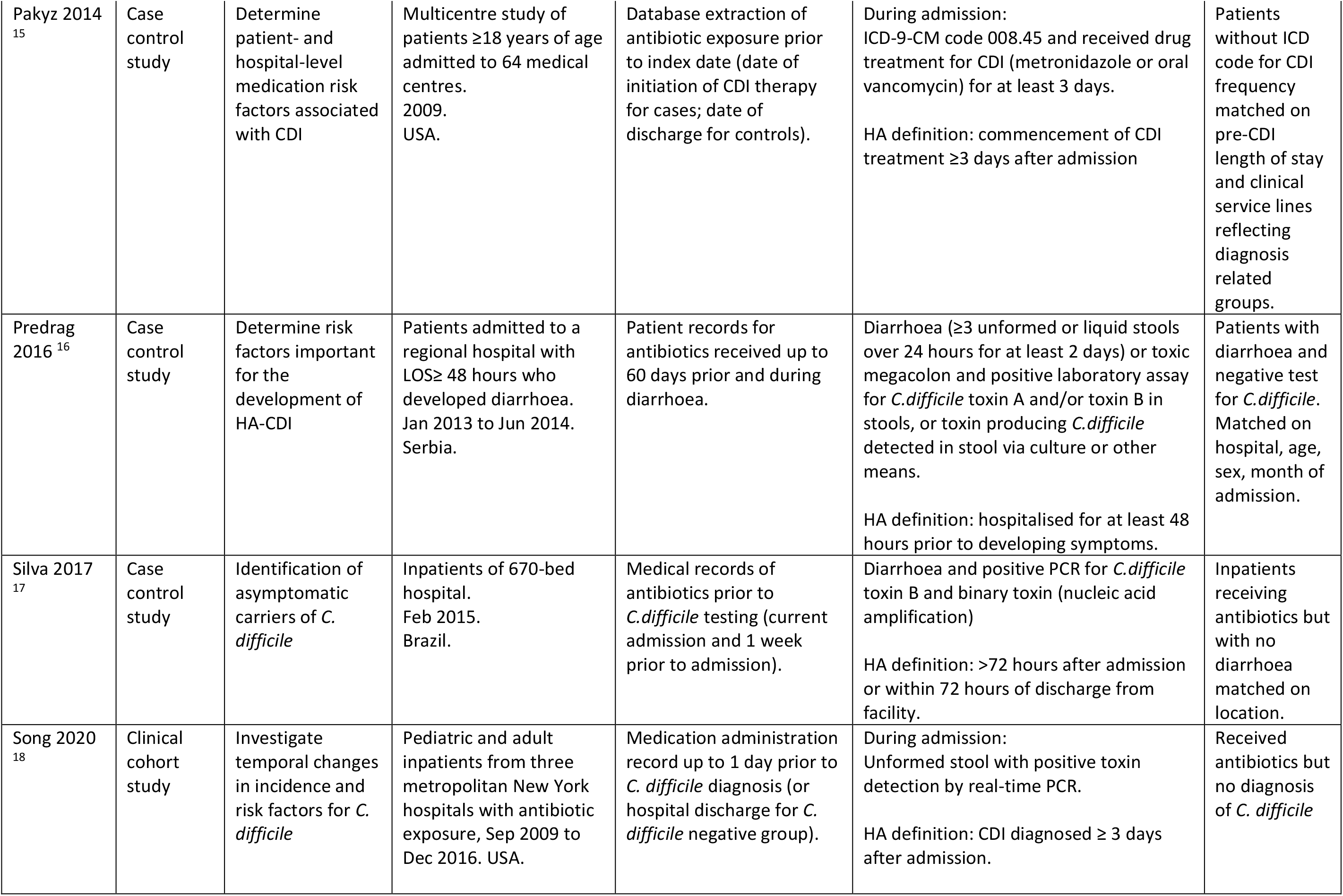
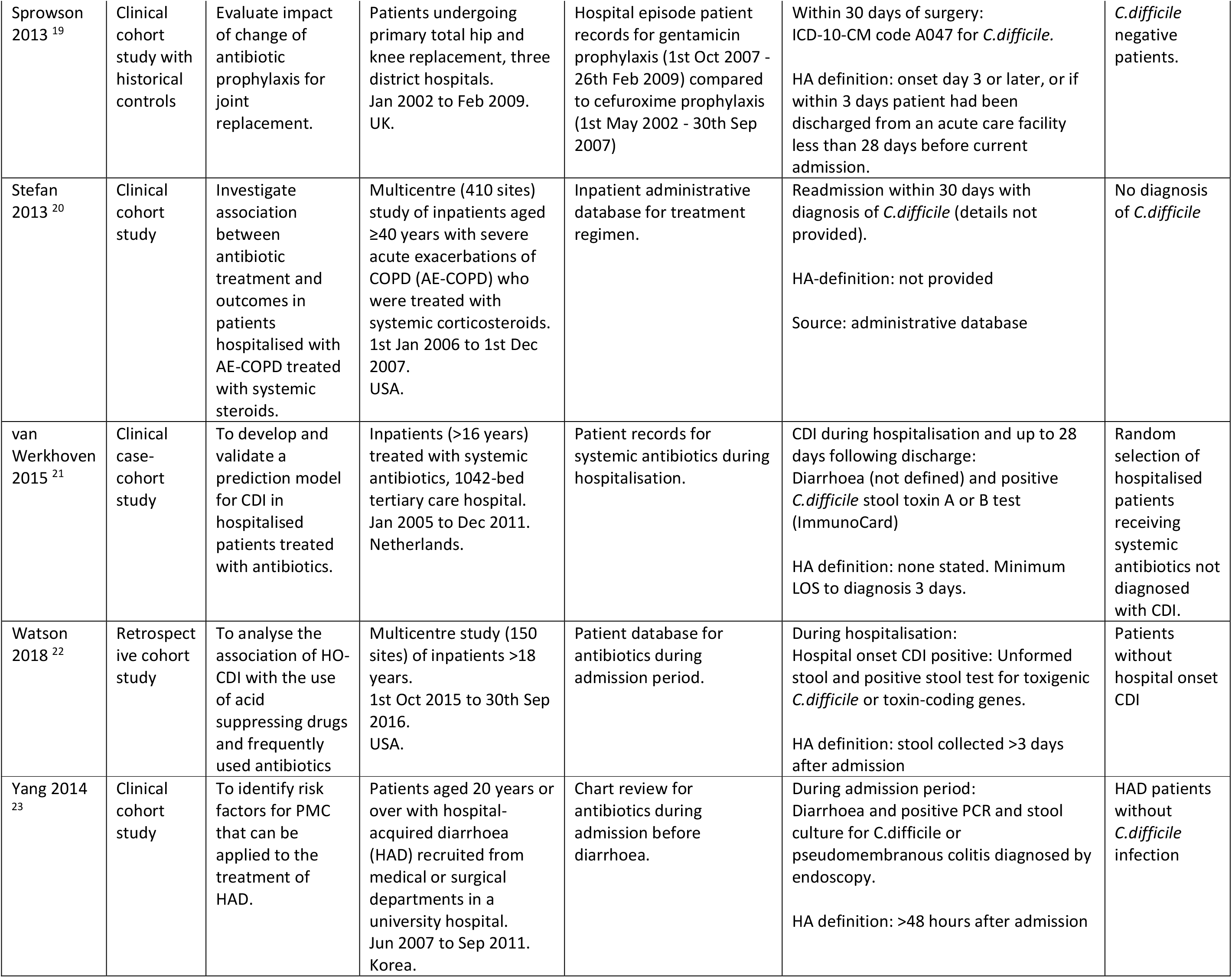
Characteristics of included studies of antibiotics and HA-CDI, 1^st^ January 2013 to 31^st^ December 2020.

| Author & year | Methods | Primary aim of study | Participants | Antibiotic exposure | Outcome |  |
| --- | --- | --- | --- | --- | --- | --- |
|  |  |  |  |  | HA-CDI case definition | Non-cases |
| Brown 2014 <sup>1</sup> | Case cohort study | CDI risk associated with duration of antibiotic therapy. | Adults (>18 years) admitted to a tertiary hospital. 1st Jun 2010 to 31st May 2012. Canada. | Pharmacy dispensing records during admission within 5 days of (i) symptom onset for CDI cases, and (ii) discharge, study termination or death for non-cases. | During hospital stay either:<br>(i) visualisation of pseudomembranes, or<br>(ii) diarrhoea (2 or more loose/watery stools in a 24-hour period with no other recognised etiology) plus positive stool PCR toxin assay for *C.difficile*.<br>HA definition: Onset >2 days after admission | 10% random sample of non-CDI cohort. |
| Cannon 2017 <sup>2</sup> | Clinical cohort study | Risk of CDI during hospitalisation for hematology-oncology patients colonised with *C.difficile*. | Admissions (LOS>1) to hematological malignancy/bone marrow transplant unit in 566-bed hospital. 12th May 2015 to 24th Sep 2015. USA. | Electronic medical record within previous 30 days | During hospital stay:<br>Diarrhoea (following IRDS/SHEA 2010 guidelines) and positive test for *C.difficile* (culture plus PCR).<br>HA definition: Diagnosis > 24h after admission | Patients that did not develop symptomatic CDI |
| Debast 2014 <sup>3</sup> | Case control outbreak study | Identify risk factors for CDI during an outbreak. | Adult inpatients of a 341-bed hospital. Apr to Sep 2005. Netherlands. | Pharmacy database records for antibiotics prescribed within and outside the hospital for preceding 3 months. | During hospital stay:<br>Diarrhoea (2 or more loose bowel movements per day) and positive stool *C.difficile* toxin EIA (Immunocard Toxin A/B, Meridien) followed by culture and PCR toxin typing.<br>HA definition: not stated; 44/45 probable HA | Patients residing at the same time and the same ward.<br>1. Non-CDI diarrhoea<br>2. Patients without diarrhoea |
| Forster 2017 <sup>4</sup> | Cohort study | To determine the potential effects of interventions on hospital-acquired CDI risk | Adult inpatients (>18 years) of a 1000-bed tertiary care hospital. 1st Mar 2004 to 1st Mar 2014. Canada. | Pharmacy database records of exposure to antibiotic class during admission. | During hospital index admission: Unformed stool sample with positive stool EIA for toxin A or B (Bio Nuclear Diagnostics).<br>HA definition: samples taken >48 hours after admission and up to 1 month after discharge. | Patients that did not develop symptomatic CDI |
| Gaertner 2015 <sup>5</sup> | Nested clinical case control study | To review and compare outcomes of patients with and without CDI infection after elective colorectal operations. | Patients undergoing elective colon or rectal surgery. Jan 2007 to Dec 2012. USA. | Nursing, anaesthesia and operative records of preoperative prophylaxis antibiotic | Up to 30 days post-operative: ICD code 008.45 from hospital database with chart confirmation of positive *C.difficile* toxin assay (PCR or EIA) and postoperative diarrhoea (≥3 loose or liquid stools per day) or high stoma output (>2000ml per day).<br>HA definition: unclear. Minimum time from surgery to CDI 2 days | Matched on age, diagnosis and procedure performed:<br>1. Diarrhoea with negative toxin assay<br>2. Patients without an assay |
| Gutierrez-Pizarra 2018 <sup>6</sup> | Clinical cohort study | To describe the incidence, clinical characteristics, and risk factors for CDI in critically ill patients. | Patients who developed diarrhoea during ICU stay (26 ICUs). 3rd Feb to 3rd Apr 2014. Spain. | Source not stated; antibiotics within 30 days prior to onset of diarrhoea. | During ICU stay: Development of diarrhoea (≥3 unformed stools within 24 hours) with positive test for *C.difficile* (method not provided).<br>HA definition: not defined. 94% of sample developed diarrhoea >48 hours after admission. | Patients with diarrhoea and negative test for *C.difficile* |
| Habayeb 2015 <sup>7</sup> | Clinical cohort with historical controls | To assess the effect of a formulary change for treatment of severe hospital-acquired pneumonia (HAP) | Inpatients with severe HAP in an acute care hospital. Jan 2011 to Jul 2012. UK. | Pharmacy records of patients treated with amoxicillin plus temocillin (Nov 2011 - Jul 2012) compared to patients treated with piperacillin/tazobactam (Jan 2011 - Nov 2011) | During inpatient treatment for severe HAP: Development of diarrhoea (at least 2 episodes of type 6 or 7 stools on Bristol stool chart in 24 hours) and positive test for *C.difficile* (stool toxin A and B detection up to 2012 followed by two-step GDH and toxin immunoassay).<br>HA definition: assumed on basis of HAP definition (> 48h after admission) | Patients that did not develop CDI |
| Hassan 2013 <sup>8</sup> | Clinical cohort | Determine the rate and characteristics of HA-CDI in subjects with type 2 diabetes admitted into acute medical wards. | Adult patients (>18 years) with type 2 diabetes consecutively admitted to acute medical wards in 700-bed tertiary hospital. Aug 2009 to May 2010. Malaysia. | Source not stated; antibiotic prescription covering at least 48 hours in the last 8 weeks prior to *C.difficile* infection. | During admission:<br>Diarrhoea (>3 loose stools within a 24 hour period for >2 days) and two positive stool Toxin A/B tests (different test kits)<br>HA definition: >48 hours after admission or <48 hours if patient hospitalised within 4 weeks prior to admission. | Patients not diagnosed with HA-CDI |
| Kirkwood 2018 <sup>9</sup> | Clinical cohort | Characterise the incidence and risks of CDI in cardiac surgery patients. | Cardiac surgery patients (≥18 years), multicentre (10 sites). Feb and Sep 2010. USA & Canada. | Source not stated; pre-operative antibiotic prophylaxis. | During 65 days postoperative follow-up: CDI diagnosis as per standard clinical practice at each site.<br>HA definition: not defined. Minimum time to CDI onset = 8 days | Patients without CDI |
| Li 2017 <sup>10</sup> | Clinical cohort | Examine the risk and disease burden of CDI amongst hospitalised HAP patients. | Patients admitted to ICU in a 3500-bed tertiary hospital with HAP who developed hospital onset diarrhoea. Jan to Dec 2014. China. | Medical records of antibiotics used for HAP therapy. | During admission:<br>Hospital onset diarrhoea (≥3 episodes within 24 hours) and detection of toxin-producing *C.difficile* by culture and PCR.<br>HA definition: ≥48 hours after hospital admission | Patients with hospital onset diarrhoea and negative for *C.difficile* |
| Lin 2015 <sup>11</sup> | Clinical cohort study | Examine risk factors for CDI amongst hospitalised patients colonised with toxigenic *C.difficile*. | Inpatients (≥18 years) with toxigenic *C.difficile* colonisation admitted to medical ward of a district general hospital. Jan 2011 to Jun 2012. Taiwan. | Medical records of antibiotics prescribed within one month before CDAD or end of follow-up. | During admission including repeat admissions (follow up until discharge, death or end of study period):<br>Diarrhoea (3 loose stools within at least a 2-day period) and detection of toxin-producing *C.difficile* by culture and PCR.<br>HA-definition: not specified. Screened for colonisation within 48 hours, follow-up for development of CDI | Asymptomatic colonised patients |
| Lopardo 2015 <sup>12</sup> | Clinical cohort study | Describe occurrence of *C.difficile*. | Inpatients at four tertiary care hospitals aged ≥40 years who received antibiotic treatment. 21st Sep 2010 to 19th Sep 2011. Mexico and Argentina. | Source not stated; antibiotics prescribed during hospitalisation prior to onset of CDI. | Follow up for 30 days after start of antibiotic therapy<br>Clinical suspicion of CDI (i.e. antibiotic-associated diarrhoea [3 or more stools in a 24 hour period]) and positive ELISA test for *C.difficile* toxins A or B and toxigenic culture (if positive on ELISA).<br>HA definition: not specified. Minimum time from admission to symptoms 3 days | Patients that did not develop CDI. |
| Maestri 2020 <sup>13</sup> | Clinical cohort study | Identify prevalence, clinical characteristics, treatment, and recurrence of CDI in patients with antimicrobial-associated diarrhoea. | Adult inpatients aged ≥18 years from eight hospitals who developed antimicrobial-associated diarrhoea. 2017-2019. Brazil. | Standardised questionnaire of antimicrobials received in 90 days preceding diarrhoea onset. | During admission; diarrhoea (≥3 liquid stools per day) and a stool test positive for toxin producing *C.difficile* by EIA or qPCR.<br>HA definition: not specified. Hospitalised >48 hours after admission. | *C.difficile* negative patients with antimicrobial-associated diarrhoea |
| Matthaiou 2019 <sup>14</sup> | Clinical cohort study | Investigate incidence of CDI in Greek ICUs and identify potential risk factors for CDI | Patients hospitalised in 3 ICU units for more than >72 hours who developed diarrhoea. 1st Jan 2014 to 31st Dec 2014. Greece. | Source not stated; antibiotics prior to and during ICU admission. | During ICU admission:<br>Diarrhoea ( ≥3 watery stools per day) and positive test for *C.difficile* (GDH and toxin A/B; toxigenic culture) and any of the following clinical signs: leukocytosis >15000 per mm<sup>3</sup>, dehydration/hypotension, shock, ileus, and megacolon.<br>HA definition - not specified. HA-CDI stated in abstract. | *C.difficile* negative patients with diarrhoea |
| Pakyz 2014<sup>15</sup> | Case control study | Determine patient- and hospital-level medication risk factors associated with CDI | Multicentre study of patients ≥18 years of age admitted to 64 medical centres. 2009. USA. | Database extraction of antibiotic exposure prior to index date (date of initiation of CDI therapy for cases; date of discharge for controls). | During admission:<br>ICD-9-CM code 008.45 and received drug treatment for CDI (metronidazole or oral vancomycin) for at least 3 days.<br>HA definition: commencement of CDI treatment ≥3 days after admission | Patients without ICD code for CDI frequency matched on pre-CDI length of stay and clinical service lines reflecting diagnosis related groups. |
| Predrag 2016<sup>16</sup> | Case control study | Determine risk factors important for the development of HA-CDI | Patients admitted to a regional hospital with LOS ≥ 48 hours who developed diarrhoea. Jan 2013 to Jun 2014. Serbia. | Patient records for antibiotics received up to 60 days prior and during diarrhoea. | Diarrhoea (≥3 unformed or liquid stools over 24 hours for at least 2 days) or toxic megacolon and positive laboratory assay for *C.difficile* toxin A and/or toxin B in stools, or toxin producing *C.difficile* detected in stool via culture or other means.<br>HA definition: hospitalised for at least 48 hours prior to developing symptoms. | Patients with diarrhoea and negative test for *C.difficile*. Matched on hospital, age, sex, month of admission. |
| Silva 2017<sup>17</sup> | Case control study | Identification of asymptomatic carriers of *C. difficile* | Inpatients of 670-bed hospital. Feb 2015. Brazil. | Medical records of antibiotics prior to *C.difficile* testing (current admission and 1 week prior to admission). | Diarrhoea and positive PCR for *C.difficile* toxin B and binary toxin (nucleic acid amplification)<br>HA definition: >72 hours after admission or within 72 hours of discharge from facility. | Inpatients receiving antibiotics but with no diarrhoea matched on location. |
| Song 2020<sup>18</sup> | Clinical cohort study | Investigate temporal changes in incidence and risk factors for *C. difficile* | Pediatric and adult inpatients from three metropolitan New York hospitals with antibiotic exposure, Sep 2009 to Dec 2016. USA. | Medication administration record up to 1 day prior to *C. difficile* diagnosis (or hospital discharge for *C. difficile* negative group). | During admission:<br>Unformed stool with positive toxin detection by real-time PCR.<br>HA definition: CDI diagnosed ≥ 3 days after admission. | Received antibiotics but no diagnosis of *C. difficile* |
| Sprowson 2013 <sup>19</sup> | Clinical cohort study with historical controls | Evaluate impact of change of antibiotic prophylaxis for joint replacement. | Patients undergoing primary total hip and knee replacement, three district hospitals. Jan 2002 to Feb 2009. UK. | Hospital episode patient records for gentamicin prophylaxis (1st Oct 2007 - 26th Feb 2009) compared to cefuroxime prophylaxis (1st May 2002 - 30th Sep 2007) | Within 30 days of surgery: ICD-10-CM code A047 for *C.difficile*.<br>HA definition: onset day 3 or later, or if within 3 days patient had been discharged from an acute care facility less than 28 days before current admission. | *C.difficile* negative patients. |
| Stefan 2013 <sup>20</sup> | Clinical cohort study | Investigate association between antibiotic treatment and outcomes in patients hospitalised with AE-COPD treated with systemic steroids. | Multicentre (410 sites) study of inpatients aged ≥40 years with severe acute exacerbations of COPD (AE-COPD) who were treated with systemic corticosteroids. 1st Jan 2006 to 1st Dec 2007. USA. | Inpatient administrative database for treatment regimen. | Readmission within 30 days with diagnosis of *C.difficile* (details not provided).<br>HA-definition: not provided<br>Source: administrative database | No diagnosis of *C.difficile* |
| van Werkhoven 2015 <sup>21</sup> | Clinical case-cohort study | To develop and validate a prediction model for CDI in hospitalised patients treated with antibiotics. | Inpatients (>16 years) treated with systemic antibiotics, 1042-bed tertiary care hospital. Jan 2005 to Dec 2011. Netherlands. | Patient records for systemic antibiotics during hospitalisation. | CDI during hospitalisation and up to 28 days following discharge: Diarrhoea (not defined) and positive *C.difficile* stool toxin A or B test (ImmunoCard)<br>HA definition: none stated. Minimum LOS to diagnosis 3 days. | Random selection of hospitalised patients receiving systemic antibiotics not diagnosed with CDI. |
| Watson 2018 <sup>22</sup> | Retrospective cohort study | To analyse the association of HO-CDI with the use of acid suppressing drugs and frequently used antibiotics | Multicentre study (150 sites) of inpatients >18 years. 1st Oct 2015 to 30th Sep 2016. USA. | Patient database for antibiotics during admission period. | During hospitalisation: Hospital onset CDI positive: Unformed stool and positive stool test for toxigenic *C.difficile* or toxin-coding genes.<br>HA definition: stool collected >3 days after admission | Patients without hospital onset CDI |
| Yang 2014<sup>23</sup> | Clinical cohort study | To identify risk factors for PMC that can be applied to the treatment of HAD. | Patients aged 20 years or over with hospital-acquired diarrhoea (HAD) recruited from medical or surgical departments in a university hospital. Jun 2007 to Sep 2011. Korea. | Chart review for antibiotics during admission before diarrhoea. | During admission period:<br>Diarrhoea and positive PCR and stool culture for *C.difficile* or pseudomembranous colitis diagnosed by endoscopy.<br>HA definition: >48 hours after admission | HAD patients without *C.difficile* infection |

**Table S2.** Summary of quality appraisal of 39 studies eligible for inclusion.

| Citation (author, publication year) | NOS quality score |  |  |  | Study design |
| --- | --- | --- | --- | --- | --- |
|  | Global score (0-9) | Selection (0-4) | Comparability (0-2) | Exposure/Outcome (0-3) |  |
| Asha 2006 | 7 | 4 | 1 | 2 | Case control |
| Baxter 2008 | 8 | 3 | 2 | 3 | Case control |
| Brown 2014 | 8 | 4 | 1 | 3 | Case cohort |
| Canon 2017 | 6 | 4 | 0 | 2 | Cohort |
| Debast 2014 | 7 | 2 | 2 | 3 | Case control |
| Dubberke 2007 | 8 | 3 | 2 | 3 | Case control |
| Forster 2017 | 9 | 4 | 2 | 3 | Cohort |
| Gaertner 2015 | 7 | 3 | 1 | 3 | Case control |
| Gutierrez-Pizarra<br>2018 | 5 | 3 | 1 | 1 | Cohort |
| Habayeb 2015 | 6 | 2 | 1 | 3 | Cohort |
| Hassan 2013 | 4 | 1 | 0 | 3 | Cohort |
| Hensgens 2012 | 8 | 4 | 1 | 3 | Case control |
| Kallen 2009 | 8 | 3 | 2 | 3 | Case control |
| Kirkwood 2018 | 7 | 3 | 2 | 2 | Cohort |
| Li 2017 | 6 | 3 | 0 | 3 | Cohort |
| Lin 2015 | 7 | 3 | 2 | 2 | Cohort |
| Loo 2005 | 7 | 4 | 2 | 1 | Case control |
| Lopardo 2015 | 5 | 3 | 0 | 2 | Cohort |
| Maestri 2020 | 6 | 4 | 0 | 2 | Cohort |
| Matthaiou 2019 | 3 | 2 | 0 | 1 | Cohort |
| McCusker 2003 | 8 | 4 | 1 | 3 | Case control |
| Minson 2007 | 5 | 4 | 1 | 0 | Case control |
| Modena 2005 | 6 | 2 | 1 | 3 | Case control |
| Muto 2005 | 8 | 3 | 2 | 3 | Case control |
| Pakyz 2014 | 9 | 4 | 2 | 3 | Case control |
| Polgreen 2007 | 7 | 3 | 1 | 3 | Case control |
| Predrag 2016 | 5 | 2 | 0 | 3 | Case control |
| Silva 2017 | 6 | 3 | 0 | 3 | Case control |
| Song 2020 | 9 | 4 | 2 | 3 | Cohort |
| Sprowson 2013 | 6 | 3 | 0 | 3 | Cohort |
| Stefan 2013 | 6 | 3 | 2 | 1 | Cohort |
| Stevens 2010 | 9 | 4 | 2 | 3 | Cohort |
| Sundram 2009 | 8 | 4 | 1 | 3 | Case control |
| Thomas 2003 | 9 | 4 | 2 | 3 | Case control |
| Valerio 2012 | 7 | 4 | 1 | 2 | Cohort |
| Van Werkhoven 2015 | 7 | 2 | 2 | 3 | Case control |
| Vesta 2005 | 8 | 4 | 1 | 3 | Case control |
| Watson 2018 | 8 | 4 | 1 | 3 | Cohort |
| Yang 2014 | 7 | 3 | 1 | 3 | Cohort |

**Table S3.**
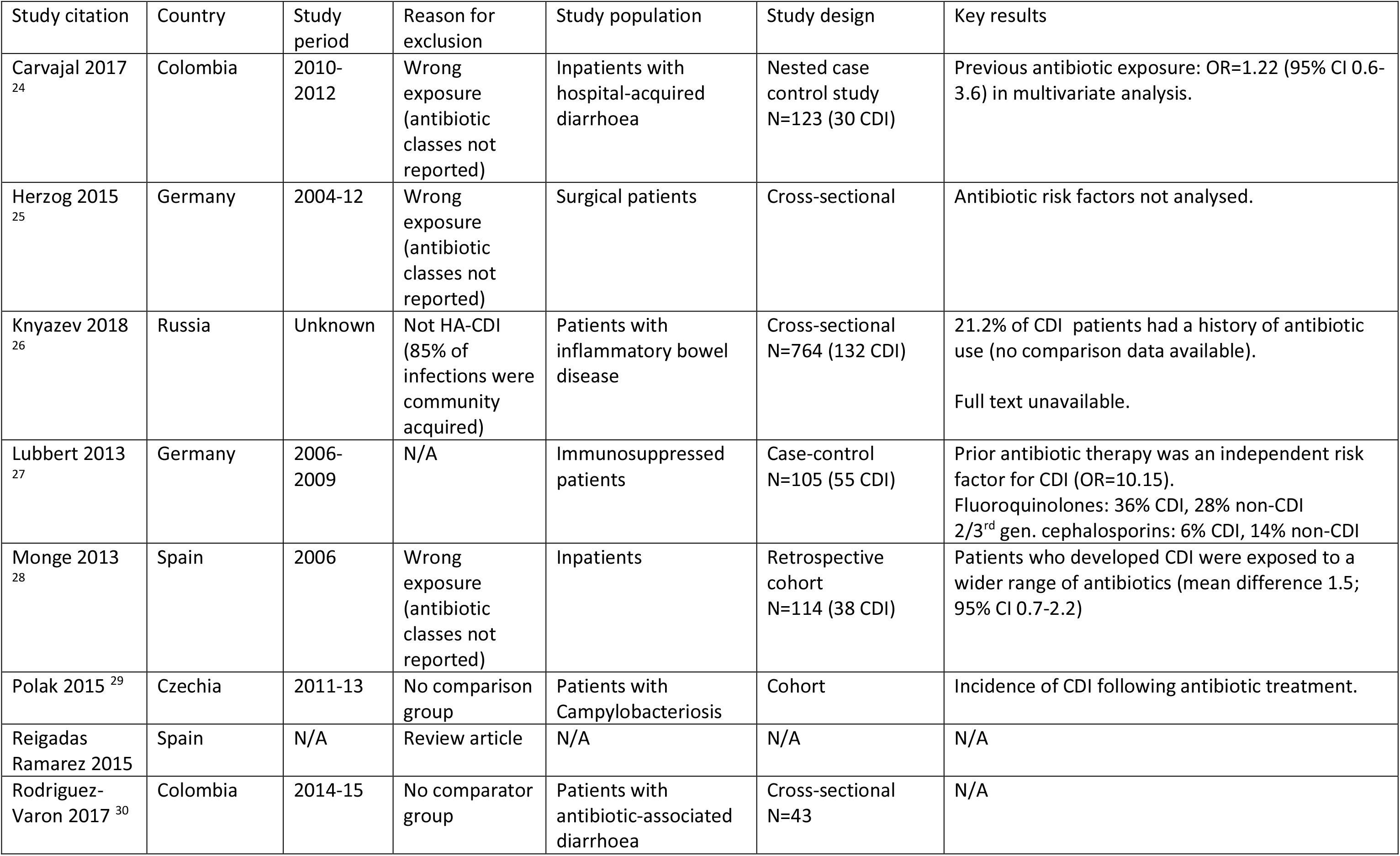
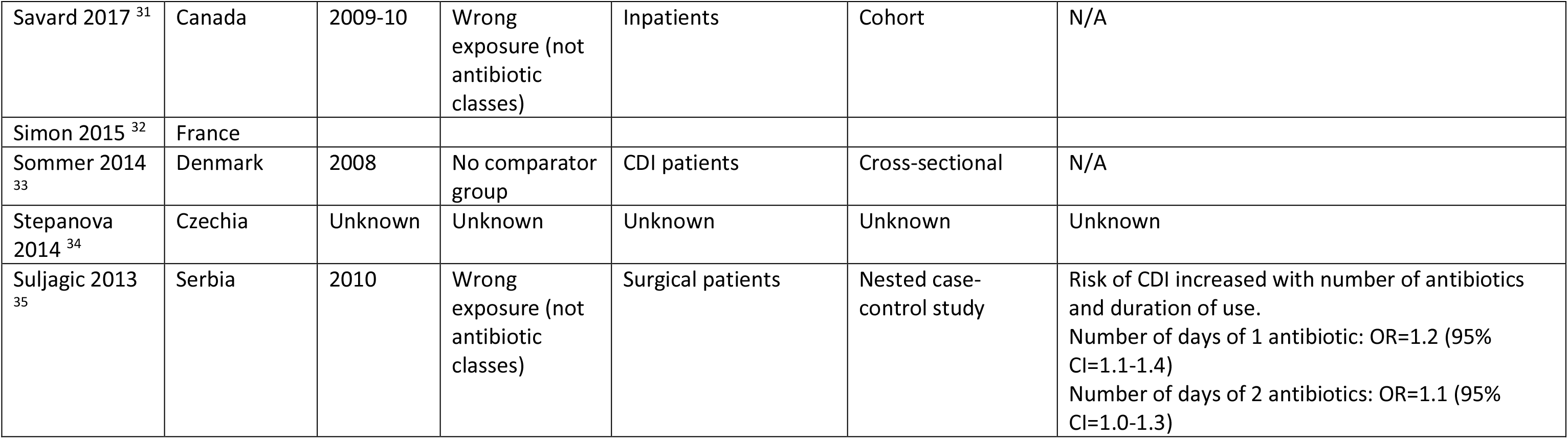
Features of eligible studies published in non-English languages.

List of studies included in review

1. Brown KA, Fisman DN, Moineddin R et al. The magnitude and duration of *Clostridium difficile* infection risk associated with antibiotic therapy: a hospital cohort study. *PLoS One* 2014; **9**: e105454.
2. Cannon CM, Musuuza JS, Barker AK et al. Risk of *Clostridium difficile i*nfection in hematology-oncology patients colonized with toxigenic *C. difficile*. *Infection Control and Hospital Epidemiology* 2017; **38**: 718-20.
3. Debast SB. *Clostridium difficile infection: the role of antibiotics in outbreak control, epidemiology and treatment*, 2014.
4. Forster AJ, Daneman N, van Walraven C. Influence of antibiotics and case exposure on hospital-acquired *Clostridium difficile* infection independent of illness severity. *Journal of Hospital Infection* 2017; **95**: 400-9.
5. Gaertner WB, Madoff RD, Mellgren A et al. Postoperative diarrhea and high ostomy output impact postoperative outcomes after elective colon and rectal operations regardless of *Clostridium difficile* infection. *Am J Surg* 2015; **210**: 759-65.
6. Gutiérrez-Pizarraya A, Martín-Villén L, Alcalá-Hernández L et al. Epidemiology and risk factors for *Clostridium difficile* infection in critically ill patients in Spain: The PROCRID study. *Enfermedades Infecciosas y Microbiologia Clinica* 2018; **36**: 218-21.
7. Habayeb H, Sajin B, Patel K et al. Amoxicillin plus temocillin as an alternative empiric therapy for the treatment of severe hospital-acquired pneumonia: results from a retrospective audit. *Eur J Clin Microbiol Infect Dis* 2015; **34**: 1693-9.
8. Hassan SA, Rahman RA, Huda N et al. Hospital-acquired *Clostridium difficile* infection among patients with type 2 diabetes mellitus in acute medical wards. *J R Coll Physicians Edinb* 2013; **43**: 103-7.
9. Kirkwood KA, Gulack BC, Iribarne A et al. A multi-institutional cohort study confirming the risks of *Clostridium difficile* infection associated with prolonged antibiotic prophylaxis. *Journal of Thoracic and Cardiovascular Surgery* 2018; **155**: 670-+.
10. Li C, Duan J, Liu S et al. Assessing the risk and disease burden of *Clostridium difficile* infection among patients with hospital-acquired pneumonia at a University Hospital in Central China. *Infection* 2017; **45**: 621-8.
11. Lin HJ, Hung YP, Liu HC et al. Risk factors for *Clostridium difficile*-associated diarrhea among hospitalized adults with fecal toxigenic *C. difficile* colonization. *J Microbiol Immunol Infect* 2015; **48**: 183-9.
12. Lopardo G, Morfin-Otero R, Moran V, II et al. Epidemiology of *Clostridium difficile:* a hospital-based descriptive study in Argentina and Mexico. *Braz J Infect Dis* 2015; **19**: 8-14.
13. Maestri AC, Raboni SM, Morales HMP et al. Multicenter study of the epidemiology of *Clostridioides difficile* infection and recurrence in southern Brazil. *Anaerobe* 2020; **64**.
14. Matthaiou DK, Delga D, Daganou M et al. Characteristics, risk factors and outcomes of *Clostridium difficile* infections in Greek Intensive Care Units. *Intensive Crit Care Nurs* 2019; **53**: 73-8.
15. Pakyz AL, Jawahar R, Wang Q et al. Medication risk factors associated with healthcare-associated *Clostridium difficile* infection: a multilevel model case-control study among 64 US academic medical centres. *J Antimicrob Chemother* 2014; **69**: 1127-31.
16. Predrag S. Analysis of risk factors and clinical manifestations associated with *Clostridium difficile* disease in Serbian hospitalized patients. *Braz J Microbiol* 2016; **47**: 902-10.
17. Silva ALO, Marra AR, Martino MDV et al. Identification of *Clostridium difficile* asymptomatic carriers in a tertiary care hospital. *Biomed Res Int* 2017; **2017**: 5450829.
18. Song J. Analyzing Risk Factors for Healthcare-Associated Infections Using Multiple Methodological Approaches. 2020.
19. Sprowson A, Symes T, Khan SK et al. Changing antibiotic prophylaxis for primary joint arthroplasty affects postoperative complication rates and bacterial spectrum. *Surgeon* 2013; **11**: 20-4.
20. Stefan MS, Rothberg MB, Shieh MS et al. Association between antibiotic treatment and outcomes in patients hospitalized with acute exacerbation of COPD treated with systemic steroids. *Chest* 2013; **143**: 82-90.
21. van Werkhoven CH, van der Tempel J, Jajou R et al. Identification of patients at high risk for *Clostridium difficile* infection: development and validation of a risk prediction model in hospitalized patients treated with antibiotics. *Clin Microbiol Infect* 2015; **21**: 786.e1-8.
22. Watson T, Hickok J, Fraker S et al. Evaluating the Risk Factors for Hospital-Onset *Clostridium difficile* Infections in a Large Healthcare System. *Clinical Infectious Diseases* 2018; **66**: 1957-9.
23. Yang BK, Do BJ, Kim EJ et al. The simple predictors of pseudomembranous colitis in patients with hospital-acquired diarrhea: a prospective observational study. *Gut and Liver* 2014; **8**: 41-8.
24. Carvajal C, Pacheco C, Jaimes F. Clinical and demographic profile and risk factors for *Clostridium difficile* infection. *Biomedica* 2017; **37**: 53-61.
25. Herzog T, Deleites C, Belyaev O et al. *Clostridium difficile* in visceral surgery. *Chirurg* 2015; **86**: 781-6.
26. Knyazev OV, Kagramanova AV, Chernova ME et al. *Clostridium difficile* in inflammatory bowel disease. *Ter Arkh* 2018; **90**: 32-6.
27. Lübbert C, Johann C, Kekulé AS et al. [Immunosuppressive treatment as a risk factor for the occurrence of clostridium difficile infection (CDI)]. *Z Gastroenterol* 2013; **51**: 1251-8.
28. Monge D, Millán I, González-Escalada A et al. The effect of *Clostridium difficile* infection on length of hospital stay. A cohort study. *Enferm Infecc Microbiol Clin* 2013; **31**: 660-4.
29. Polák P, Vrba M, Bortlíček Z et al. Campylobacteriosis at the Department of Infectious Diseases of the University Hospital Brno in 2011-2013: a retrospective study. *Epidemiol Mikrobiol Imunol* 2015; **64**: 153-9.
30. Reigadas RamÃ•rez E, Bouza Santiago E, Universidad Complutense de M et al. Estudio de la infección “*Clostridium difficile*”: Incidencia, epidemiología, características clínicas, factores de riesgo de gravedad y recurrencia. 2015.
31. Savard Aa, Brisson M. Impact des caractéristiques des hôpitaux québécois sur le taux d’incidence d’infection à *Clostridium difficile* 2017.
32. Simon E, Cherfaoui T. La réévaluation des traitements par inhibiteur de la pompe à proton chez les patients traités au long cours en médecine générale dans l’Ille et Vilaine. Université de Rennes 1 & Université européenne de Bretagne., 2015.
33. Sommer TN, Ravn P, Gjørup I. *Clostridium difficile* ribotype 027 is a challenge. *Ugeskr Laeger* 2014; **176**.
34. Štěpánová J, Tomášková H. Epidemiology of infection caused by *Clostridium difficile Hygiena* 2014; **59**: 131-9.
35. Šuljagić V, Djordjević D, Lazić S et al. Epidemiological characteristics of nosocomial diarrhea caused by *Clostridium difficile* in a tertiary level hospital in Serbia. *Srp Arh Celok Lek* 2013; **141**: 482-9.

**Figure S1.**
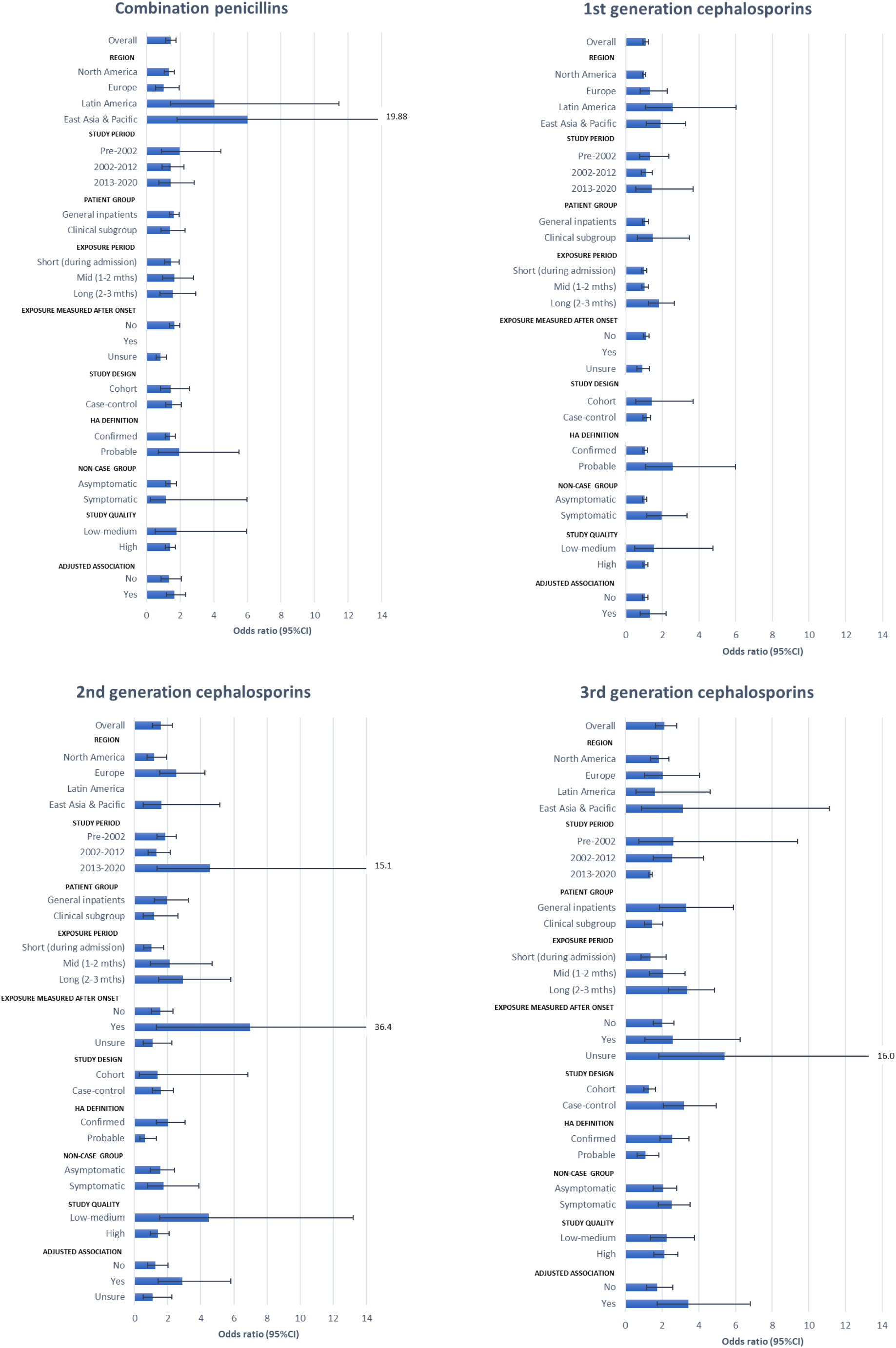
Sub-group analysis – combination penicillins and cephalosporin sub-classes.

**Figure S2.**
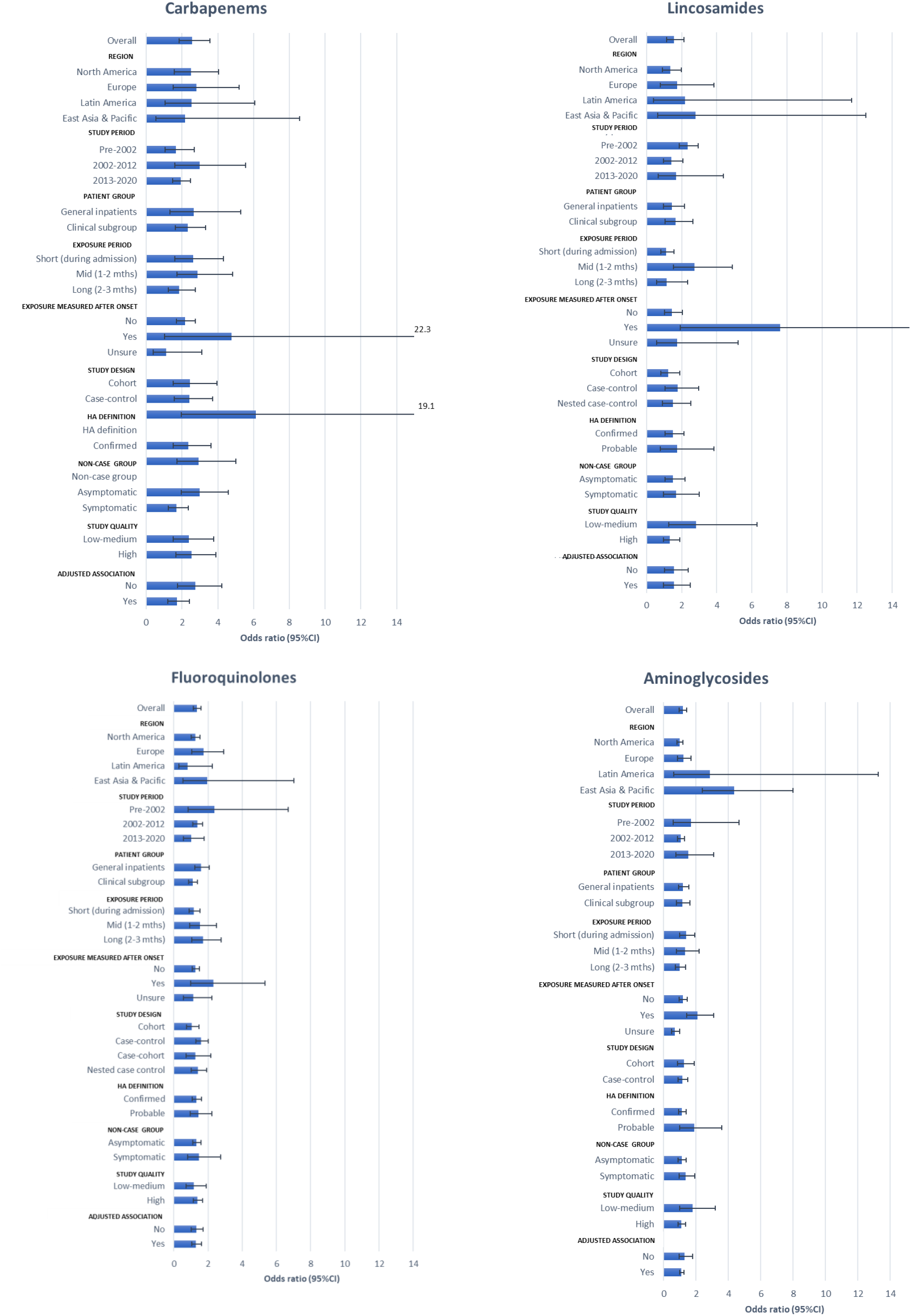
Sub-group analysis – carbapenems, lincosamides, fluoroquinolones, aminoglycosides.

**Figure S3.**
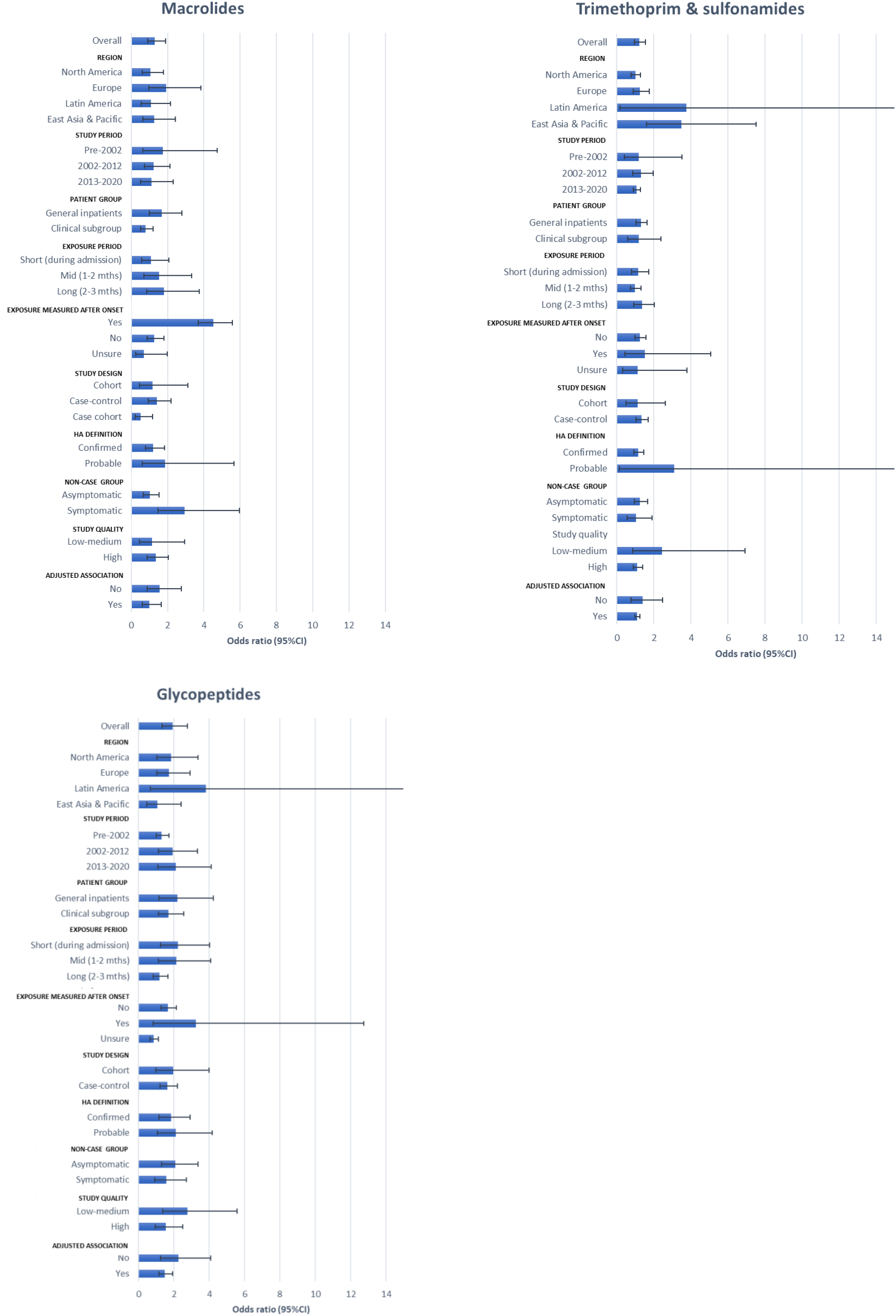
Sub-group analysis – macrolides, trimethoprim-sulfonamides and glycopeptides.

**Figure S4.**
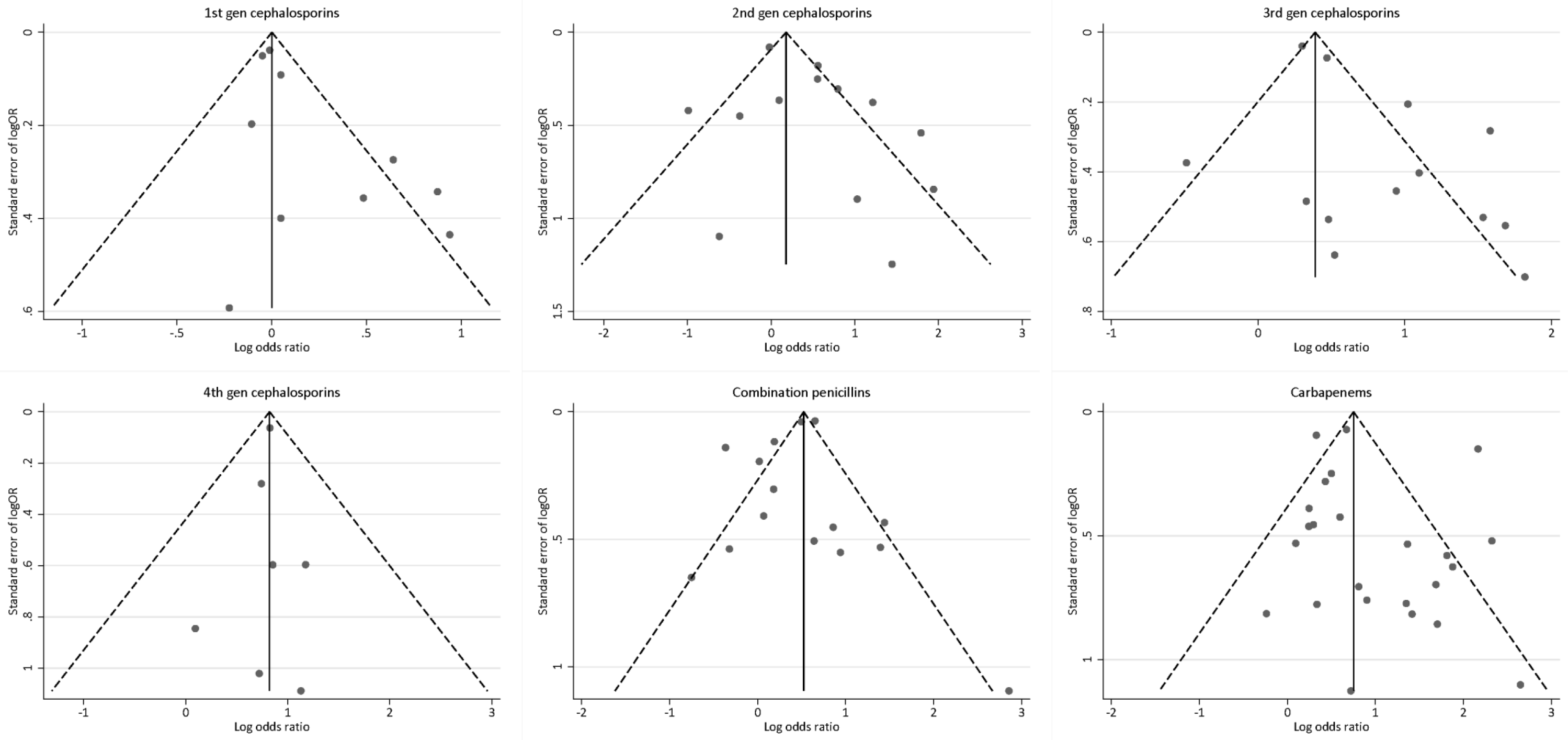
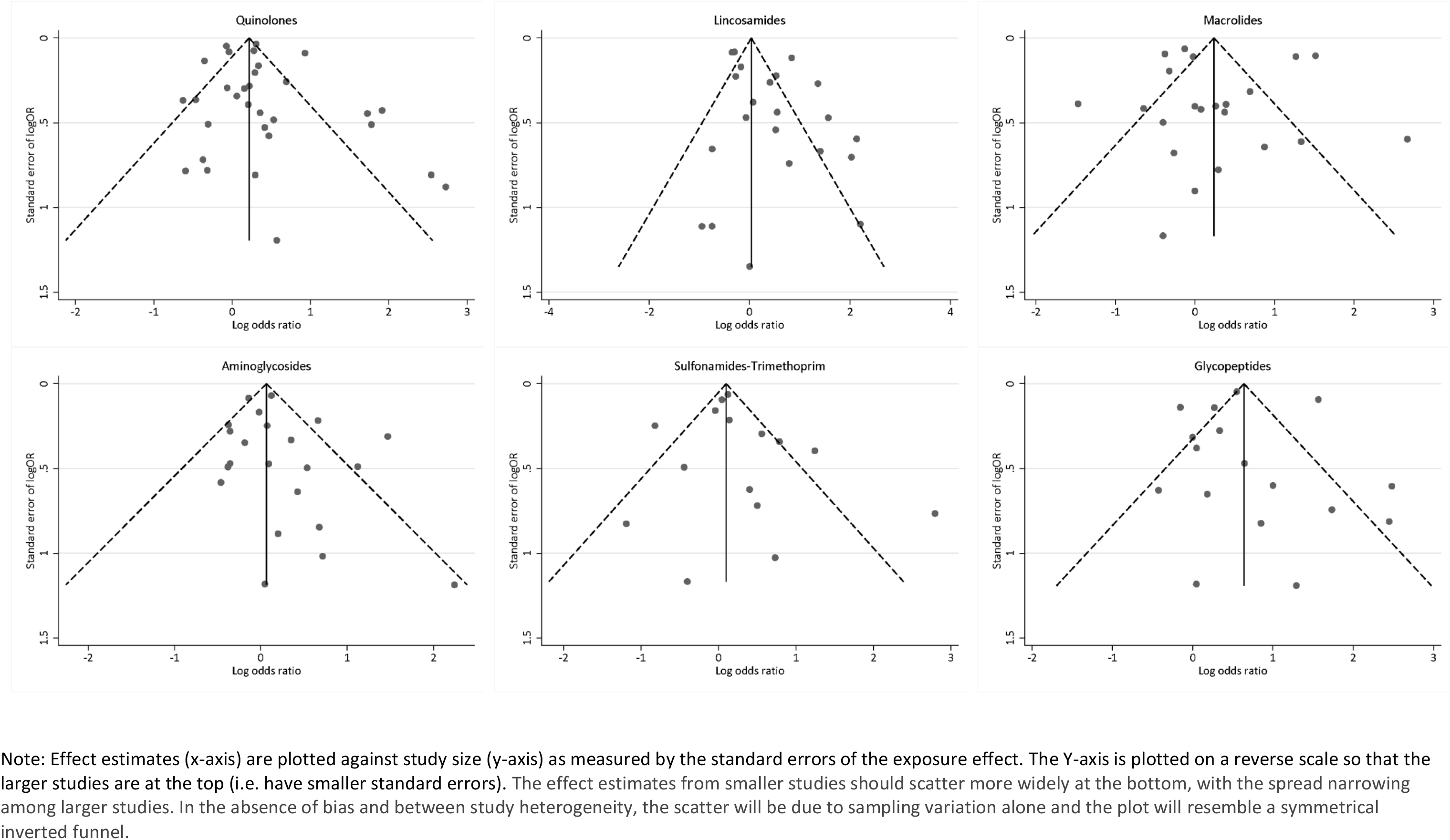
Funnel plots.

## Notes

Transparency declarations: All authors have no conflicts to declare in relation to this work.

### Competing Interest Statement

The authors have declared no competing interest.

### Clinical Protocols

https://www.crd.york.ac.uk/prospero/display_record.php?ID=CRD42020181817

### Funding Statement

This study did not receive any external funding.

### Author Declarations

The study is a systematic review and meta-analysis of peer reviewed studies and is therefore exempt from ethics committee approval.

